# Personalized Autoantibody Profiling Distinguishes Early-stage Breast Cancer from Benign Disease

**DOI:** 10.64898/2026.01.15.26344214

**Authors:** Kathryn A Lyon, Justin C Rolando, David R Walt

## Abstract

**Background:** Early and accurate detection of breast cancer and differentiation from benign breast disease remains a substantial challenge, with about 70% of diagnostic breast biopsies having no malignant findings. Tumor-associated Autoantibodies represent the immune system’s response to a neoplasm and are a promising biomarker group for the early diagnosis of breast cancer by liquid biopsy.

**Methods:** In this study, we quantified the IgM and IgG titers to 525 Tumor Associated Antigens in a prospectively-collected cohort of 50 serum samples from donors with benign breast disease and donors with early-stage breast cancer. The considerable number of antibodies analyzed enabled us to account for variations in individual immune profiles through *z-*score normalization of each donor’s total antibody distribution. Differentially expressed antibodies were identified using Mann-Whitney *U* tests (*p* < 0.05) and fold-change analysis (fold-change > ± 1.2). For each donor, we calculated the total number of “high-titer” antibodies, defined as antibodies with relative concentrations > 3 SD above the cohort mean. Logistic regression classifiers were then built using differentially expressed biomarkers and high-titer antibody counts to distinguish benign breast disease from breast cancer.

**Results:** We identified 25 differentially expressed antibodies between the benign and cancer groups. A down-selected panel of eight antibodies demonstrated good performance in a logistic regression classifier to distinguish benign disease from invasive carcinomas (AUC-ROC = 0.83 ± 0.14). High-titer antibody analysis revealed that the benign group had a higher prevalence of donors with elevated IgG immune response, and donors displayed antibody signatures unique to their individual disease pathway.

**Conclusions:** This study identifies an eight-antibody panel with promising diagnostic potential to distinguish benign breast disease from early-stage breast cancer. The z-score normalization approach and analysis of individual donors’ high-titer antibody profiles represent a novel approach towards personalized cancer immunology. This study provides encouraging preliminary evidence supporting the promise of tumor-associated autoantibody profiling for distinguishing benign and malignant breast disease, warranting future studies in larger cohorts.

## Introduction

Despite remarkable improvements in the field of breast cancer diagnostics, the accurate distinction of benign disease from early-stage cancer remains a substantial challenge. The implementation of mammography as a screening technique paired with new treatment approaches has increased the five-year survival rate of breast cancer by 16% since 1975^1^. However, about ten percent of mammographic screenings lead to additional diagnostic procedures in patients without malignant disease^2,3^. In fact, about 70% of patients who are recommended to receive a diagnostic biopsy have benign breast disease, with only about 30% of biopsies resulting in a cancer diagnosis^4^. The high-false positive rate underscores the need for complementary molecular diagnostics capable of distinguishing benign from malignant lesions following abnormal image findings.

Liquid biopsy is a promising companion diagnostic modality to mammography that could substantially reduce the percentage of patients who receive a core needle biopsy without a malignant finding. In a liquid biopsy, a blood sample is collected, and circulating markers associated with the tumor, or the biological response to the tumor, are quantified^5–7^. Tumor-associated antigens (TAAs) in the peripheral blood have been heavily researched as diagnostic biomarkers for breast cancer, but there has been limited success finding biomarkers with adequate sensitivity and specificity to distinguish early invasive disease from benign breast disease. Even “conventional” breast cancer biomarkers such as CA15-3, CEA, CA27.29, He4, and CA125, while capable of distinguishing advanced disease from normal donors, lack the sensitivity and specificity needed for early-stage detection^8–10^.

Antibodies that arise from the immune response to TAAs have been hypothesized to hold high sensitivity and specificity for early-stage cancers that has been lacking from other classes of biomarkers^11–13^. Driven by the substantial need for a more accurate breast cancer screening test, many prior studies of TAAbs have focused on the distinction of invasive breast disease from a normal patient class^14–17^. This focus overlooks the clinically relevant, yet more diagnostically challenging scenario associated with a radiological image finding of distinguishing between malignant and benign breast disease. Few studies have investigated the use of tumor-associated autoantibodies (TAAbs) to differentiate breast cancer from benign breast disease, and those that have are limited by differences in pre-analytical factors, such as the use of different sample collections between diagnostic classes^18,19^. Additionally, none of these studies measured a sufficiently large number of TAAbs to account for variations in immune activity between patients and relative autoantibody titers. Thus, the role of TAAbs in distinguishing early-stage breast cancer from benign breast disease remains unresolved.

We hypothesize that both the concentration and pattern of TAAbs differ in early-stage breast cancer patients compared to benign breast disease. Since the immune system likely encounters and eliminates nascent tumors multiple times before malignancy develops, TAAbs may serve as a molecular record of prior antigenic exposures or act as oncogenic drivers (**Fig. 1**). In our specific study, all donors had neoplasms at the time of sample collection, and thus, we propose that both individuals with benign disease and those with early-stage breast cancer would have high TAAb titers, reflective of the persistent antigenic exposure from a neoplasm. However, we expect that the antigen recognition pattern will differ between diagnostic groups as a result of distinct immune responses to benign versus malignant pathology. In contrast, while healthy individuals may also retain TAAbs from previously cleared nascent tumors, these antibodies should be present at lower titers in the absence of active disease or directed against fewer antigens. Thus, by comparing patients with active neoplasms, we aim to identify distinct disease-specific antibody signatures rather than simply measuring residual immune memory.

**Figure 1:**
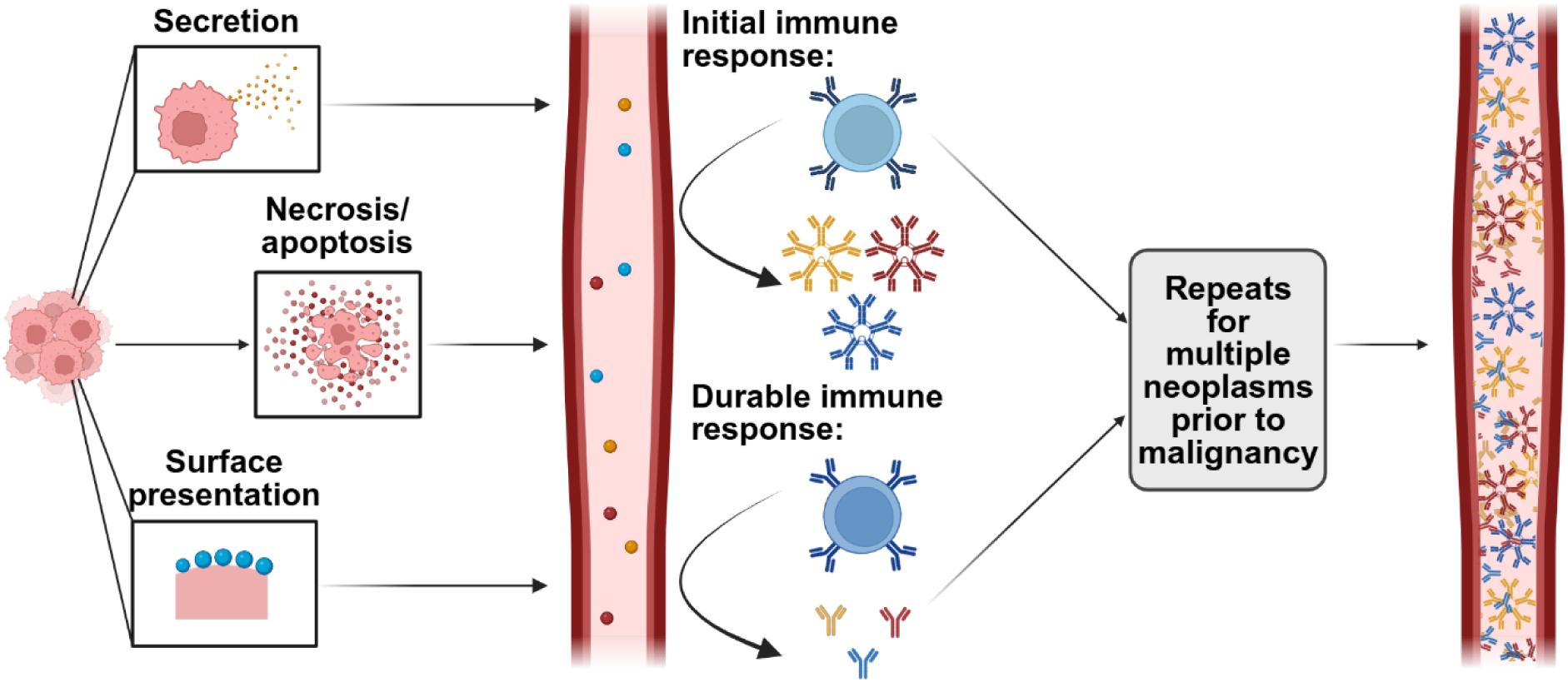
Schematic representation of immune activation and amplification in response to neoplastic growth. Disease-specific antigens are released through secretion, necrosis or apoptosis, or surface presentation. In response to these antigens, B-cells become activated, differentiate into plasma cells, and initially produce IgM. With further exposure to the antigen and stimulation from T-cells, the B-cells undergo class switch recombination, resulting in IgG production. This response is amplified by exposure to multiple neoplasms, resulting in the presence of both IgM and IgG TAAbs in the peripheral blood. Created in BioRender. (2025) https://BioRender.com/0dcdvly

Because nascent tumors express antigens in low abundance, direct detection of TAAs can be challenging. In contrast, TAAbs are biologically amplified by the immune system, circulating at higher concentrations than TAAs, and are therefore more readily detectable in the peripheral blood^20^. Moreover, TAAbs have longer circulating half-lives than TAAs, and thus can be detected long after the initial immunogen has been cleared. Importantly, TAAb production has been observed well in advance of the onset of clinical symptoms, supporting their potential as a promising biomarker class for distinguishing early-stage breast cancer from benign breast disease^21–23^.

In this study, we conducted an extensive screening of TAAbs in serum samples from 26 donors with breast cancer and 24 donors with benign breast disease. Using the Sengenics i-Ome Cancer Array, we quantified immune responses to a panel of 525 TAAs selected through an extensive literature search for autoantigens with oncological relevance, measuring both the IgM and IgG titers for each antigen (1,050 total measurements). This study represents one of the first broad screening efforts of TAAbs specifically focused on differentiating early-stage breast cancer from benign breast disease. We observed that, not unexpectedly, each donor mounts a distinct antibody response to their specific disease, with some donors exhibiting hyperactive or hypoactive responses. This indicates that immune activation is highly individualized, necessitating the normalization of individual TAAb distributions for meaningful comparison—an approach essential for broad diagnostic applicability. Interestingly, we found that the number of benign donors with an activated IgG immune response, characterized by many high-expression antibodies, was greater than the number of cancer donors with an activated IgG immune response and that many identified differentially expressed biomarkers were elevated in the benign group, indicating a potential protective role for some TAAbs. While no single biomarker was sufficient to distinguish between diagnostic classes, combining multiple TAAbs into a classifier enabled us to distinguish between benign and early-stage breast cancer. These findings support the potential of TAAb signatures as diagnostic tools to improve early detection and risk stratification in breast cancer.

## Methods

### Sample Collection and Cohort Description

Blood samples were prospectively collected between July 2022 and December 2023 at Brigham and Women’s Faulkner Hospital. Samples were obtained at the time of a participant’s screening or diagnostic mammogram, or prior to an image-guided biopsy or localization procedure. Written informed consent was obtained from all donors in accordance with Mass General Brigham Institutional Review Board Protocol # 2022P000451. All samples were processed within two hours of collection: blood was centrifuged twice at 2,600 rcf and 4 °C for 15 minutes, aliquoted into 0.5 mL volumes, and stored at −80 °C until analysis.

Most samples in the cancer group (n = 26) were collected prior to breast cancer diagnosis and represented the donor’s first incidence of cancer. However, two donors were receiving chemotherapy treatments at the time of the sample collection. In the benign sample group (n = 24), two donors converted to breast cancer within the two-year period after sample collection. Additionally, two donors had prior incidences of cancer: one donor had a history of breast cancer seven years prior and had undergone previous radical mastectomy, and one donor had a history of skin cancer one year prior (this donor was also one of the two donors that converted to breast cancer within two years after sample collection). Neither donor with a prior malignancy was receiving treatment at the time of collection, and both were in complete remission from their previous malignancy.

### Sample and Quality Control Analysis

The i-Ome Cancer Array (Sengenics, Malaysia) was used to screen for both IgM and IgG responses to a panel of 525 TAAs **(SI Table 1)**. This platform utilizes KREX Technology to perform high throughput, multiplexed antibody profiling with a panel of cancer-related antigens^24,25^. The antigens in this panel consist of a diverse set of tumor-associated proteins, including conventional TAAs (*e.g.*, TP53^26^, ENO1^27^, HER2^28^), cancer-testis antigens (*e.g.*, MAGE^29^, GAGE^30^, and SSX^31^ protein families), oncogenic and tumor suppressor proteins (*e.g.*, BRAF^32^, CDK4^33^, TP63^34^), cytokines and immune regulators (*e.g.*, IL6^35^, CXCL10^36^, TNFα^37^), metabolic and stress-related proteins (*e.g.*, HSP90^38^, PRDX6^39^, MIF^40^), developmental markers (*e.g.*, NANOG^41^, POU5F1^42^, SOX2^43^), and structural proteins (*e.g.*, COL1A1^44^, THBS1^45^, VIM^46^). Relative Fluorescent Intensity (RFI) was used to quantify the concentration of each antibody, with units calculated as the difference between the foreground signal intensity and background signal intensity of each spot. Each antibody was quantified in technical quadruplicate and the RFIs for replicate spots were averaged to calculate the mean RFI for each antigen in each sample.

Multiple quality controls were implemented to ensure the validity of the measurements. To assess the binding capacity of the secondary antibody, IgG and IgM were diluted in a 6-step 2-fold serial dilution starting from 12.5 µg/mL (**SI Fig. 1)**. Linearity of detection was confirmed with average R^2^ values of 0.99 for both IgM and IgG. A constant concentration of Cy5-labeled BSA was included for each array on each slide as a housekeeping probe to account for variation in the printing process between arrays and slides. Arrays passed quality control if the CV of the Cy5-BSA control was ≤ 15%.

Six empty vector controls were included on each array. Two of these serve as positive controls for the anti-cMyc assay to verify antigen print success, two are used to verify that antibodies are only binding to the antigens rather than the biotin carboxyl carrier protein, and two are empty control vectors for insect cell expression to verify that antibodies are binding to the antigens rather than vectors in expression lysates. Samples were also tested for the presence of Poly-Specific Antibodies (PSA) to evaluate the non-specific binding capacity of each sample. In the IgM panel, three samples were PSA positive (one benign, two cancer), and in the IgG panel, twelve samples were PSA positive (six benign, six cancer).

For each antigen with a CV ≥ 20% (n = 4 technical replicates), the spot contributing most to the high CV across replicates was excluded and the overall CV was recalculated. Antigens with CV ≥ 20% and fewer than three technical replicates remaining were flagged as having a high CV and excluded from further analysis. Samples with more than 1% of detectable antigens flagged as high CV were also excluded. Additionally, three buffer-only controls were included to establish background intensity and confirm the antigen-specific signals were above the background.

### Statistical Analyses

The mean RFI values were log_2_ transformed to reduce distributional skew and stabilize variance between antibodies with different expression levels. To account for variation in signal intensity between arrays, data were normalized to Net Intensity (NetI) using the array-Loess normalization^47^. Five pooled normal samples were included on each array to validate the array-Loess normalization procedure using a pair-wise correlation as a performance metric **(SI Fig 2).** A strong correlation indicated that the normalization did not impact the observed TAAb concentration between arrays. Following analysis of the NetI data, a *z*-score normalization was applied to the biomarker distribution of each donor. This approach centers the means of all donors’ IgM and IgG distributions and equalizes their standard deviations.

A two-tailed Mann-Whitney *U* Test was performed on each IgM and IgG TAAb to identify those differentially expressed in the serum of breast cancer donors compared to individuals with benign breast disease. For each TAAb, the fold-change of the raw or normalized NetI values between diagnostic classes was also calculated. TAAbs were included in further evaluation if *p*-value < 0.05, with a fold-change threshold of ≥ ± 1.2. TAAbs not meeting these criteria were excluded from downstream analysis.

A covariate analysis was performed to identify potential confounding variables. Ordinary least squares (OLS) multiple linear regression (MLR) was performed independently for each biomarker using the Statsmodels package v0.14 in Python v3.12. For each model, biomarker concentration was regressed against covariates (age, tumor size, and tumor grade). Model performance was evaluated using standard regression metrics, including R^2^, R^2^ adjusted for the number of variables, F-statistic and its associated significance. Missing values were imputed with the median prior to analysis. To evaluate multicollinearity, the variance inflation factor (VIF) was calculated for each covariate; with VIF > 10 indicating problematic multicollinearity.

Classifiers were developed to distinguish benign from cancer samples using a logistic regression estimator with the liblinear solver, L2 regularization, and default inverse regularization strength (C = 1). To identify the most informative biomarkers for classification, we implemented Recursive Feature Elimination with Cross-Validation (RFECV) using a nested cross-validation framework with five-fold StratifiedKFold in scikit-learn (v1.6). The outer loop preserved class balance across folds to provide unbiased performance estimates, while the inner loop optimized the number of selected features by maximizing the mean area under the curve of the receiver operating characteristic (AUC-ROC). Confidence intervals for AUC-ROC were computed using 10 repetitions of five-fold cross-validation to account for variability from random fold assignment given the limited sample size. Model interpretability was assessed using SHAP (SHapley Additive exPlanations, v0.49) with the LinearExplainer to estimate feature specific contributions to model predictions.

All modeling was performed within a single discovery cohort, and no independent validation set was available. The reported performance metrics therefore represent internal cross-validation estimates rather than performance generalizability. The reduced TAAb panel identified here should be considered a set of candidate biomarkers requiring confirmation in an independent cohort and, ideally, in an orthogonal assay format. This study represents a biomarker discovery effort, and the findings require prospective validation before clinical application.

## Results

### Cohort Characteristics

Blood serum samples from 24 donors with benign breast disease and 26 donors with breast cancer were analyzed for their IgM and IgG responses to 525 TAAs. Two samples in both the IgM and IgG panel failed quality control due to a high CV for > 1% of detectable antigens and were excluded from further analysis. In the IgM panel, both excluded samples were in the cancer group (**Table 1**). In the IgG panel, one sample was excluded from each group. The excluded cancer sample was from a donor receiving chemotherapy at the time of sample collection (one of two), while the excluded benign sample was from a donor who developed breast cancer two years after sample collection.

**Table 1:**
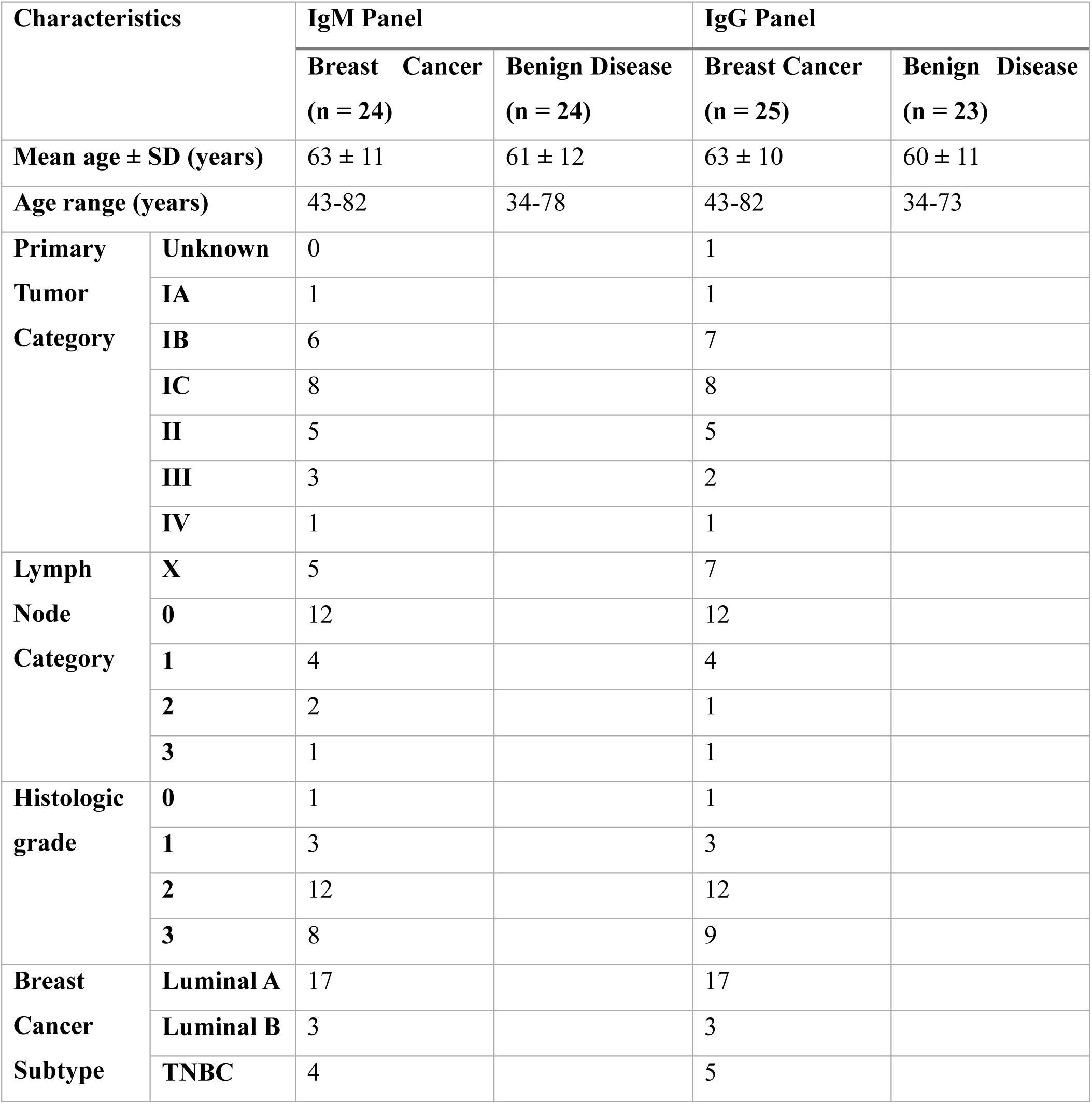
Donor Characteristics.

| Characteristics |  | IgM Panel |  | IgG Panel |  |
| --- | --- | --- | --- | --- | --- |
|  |  | Breast Cancer<br>(n = 24) | Benign Disease<br>(n = 24) | Breast Cancer<br>(n = 25) | Benign Disease<br>(n = 23) |
| Mean age ± SD (years) | | 63 ± 11 | 61 ± 12 | 63 ± 10 | 60 ± 11 |
| Age range (years) |  | 43-82 | 34-78 | 43-82 | 34-73 |
| Primary Tumor Category | Unknown | 0 |  | 1 |  |
|  | IA | 1 |  | 1 |  |
|  | IB | 6 |  | 7 |  |
|  | IC | 8 |  | 8 |  |
|  | II | 5 |  | 5 |  |
|  | III | 3 |  | 2 |  |
|  | IV | 1 |  | 1 |  |
| Lymph Node Category | X | 5 |  | 7 |  |
|  | 0 | 12 |  | 12 |  |
|  | 1 | 4 |  | 4 |  |
|  | 2 | 2 |  | 1 |  |
|  | 3 | 1 |  | 1 |  |
| Histologic grade | 0 | 1 |  | 1 |  |
|  | 1 | 3 |  | 3 |  |
|  | 2 | 12 |  | 12 |  |
|  | 3 | 8 |  | 9 |  |
| Breast Cancer Subtype | Luminal A | 17 |  | 17 |  |
|  | Luminal B | 3 |  | 3 |  |
|  | TNBC | 4 |  | 5 |  |

For donors with available TNM staging, the majority had early-stage disease. In both the IgM and IgG panels, 11 donors were Stage I and four were Stage II, with fewer donors classified as late-stage disease (IgM: n = 4 Stage III; IgG: n = 3 Stage III). There were five donors in the IgM panel and seven in the IgG panel for whom regional lymph nodes were not assessed, and therefore TNM staging was not available; however, all of these donors had tumors classified as T0 or T1, indicating they would likely be defined as Stage I or II disease, although a definitive staging cannot be made without regional lymph node assessment. The breast cancer group was assembled to reflect the heterogeneity of multiple breast cancer subtypes to the extent possible with the samples available in our collection. Similarly, the benign cohort was composed of donors with diverse benign findings including fibroadenomas, hyperplasia, and cysts.

### NetI data identifies limited differences in antibody response between benign and cancer donors

We first aimed to identify TAAbs that were differentially expressed between individual donors and analyzed trends within the preliminary data set. We cleaned the IgM and IgG datasets by removing outliers with high CVs. We then generated heatmaps of the NetI of each IgM (**SI Fig. 3a)** and IgG (**SI Fig. 3b)** antibody across all donors to visualize relative expression levels. Both donors and antibodies were clustered hierarchically with average linkage based on Pearson correlation. This analysis identified groups of donors that exhibited similar trends in their antibody distributions, though the donors did not cluster by diagnosis or cancer subtype for either panel. Additionally, some donors expressed individual TAAbs at high relative concentrations, which were not conserved between donors, indicating that most donors had a unique response to their specific disease.

We next identified antibodies that were differentially expressed between the benign and cancer groups in the NetI data set. A two-tailed Mann-Whitney *U* Test and fold-change analysis were used to compare the IgM (**SI Table 2**) and IgG (**SI Table 3**) expression levels between diagnostic classes, with significance defined as *p* < 0.05 and fold-change > ± 1.2. This analysis identified one differentially expressed IgM biomarker, anti-RHOX Homeobox Family Member 2 (RHOXF2; **Fig. 2a**), and three differentially expressed IgG biomarkers: anti-Pro-Caspase-8 (Pro-CASP8), anti-Collagen Type I Alpha 1 Chain (COL1A1), and anti-SAM Pointed Domain Containing ETS Transcription Factor (SPDEF) (**Fig. 2b**). The *p-*values and the fold-changes for each of the differentially expressed biomarkers are reported in **Table 2**.

**Figure 2:**
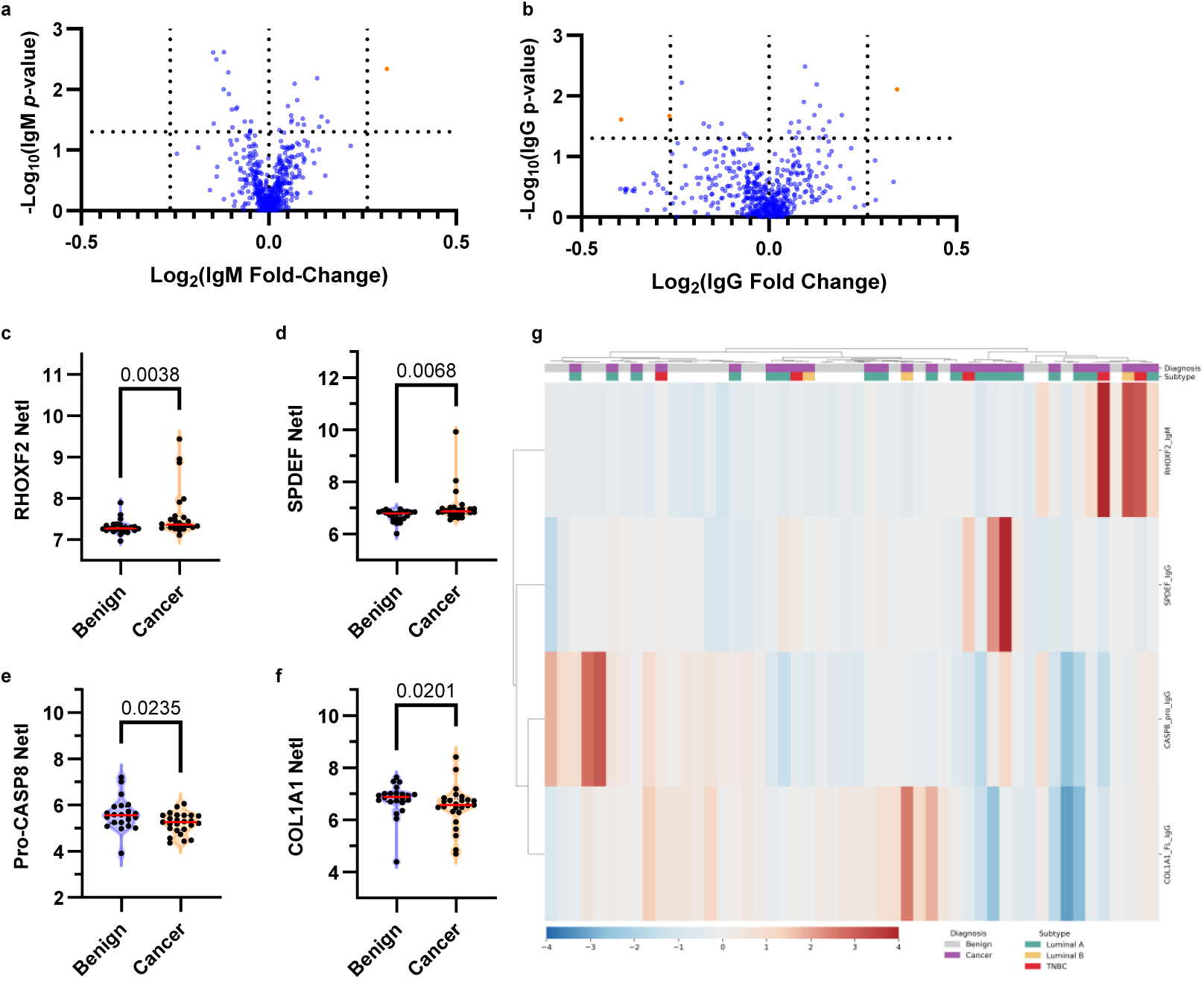
Identification of differentially expressed IgM and IgG biomarkers between benign and breast cancer donors in the NetI data set. The volcano plots show the −log_10_(*p*-value) and log_2_(fold-change) for each biomarker measured for each: **a)** the IgM response and **b)** the IgG response to 525 TAAs. Each point represents the mean of n = 4 technical replicates (n = 48 samples). The horizontal dashed line indicates statistical significance (*p*-value of 0.05). Vertical dashes indicate a fold-change > ± 1.2. Points to the left of the leftmost vertical line are biomarkers with a higher mean signal in the benign group than the cancer group, whereas points to the right of the rightmost vertical dashed line are biomarkers with higher mean signal in the cancer group. Orange points indicate biomarkers that meet both *p*-value and fold-change significance criteria. Each violin plot shows the distribution of the NetI values of each sample, distinguished by diagnostic class, for each IgM (**c**) and IgG (**d-f**) antigen identified as significant (*p* < 0.05, fold-change > ± 1.2). Each point represents the mean value of four technical replicates, or two to three if any replicates were removed for a high CV (> 20%). The *p*-values were calculated using a Mann-Whitney *U* test. The red bar shows the median. **g)** A heat map of the relative expression of the identified significant biomarkers (rows) across all subjects (columns). Red represents high relative expression while blue represents low relative expression. Both donor and biomarker axes are clustered hierarchically using average linkage based on Pearson correlation. The diagnosis and subtype of each donor are identified in the top two rows of the plot.

**Table 2:** Summary of significant TAAbs identified by differential expression analysis of raw data. Displayed are the antigens to which the TAAbs are differentially expressed that meet both the *p*-value and fold-change threshold for significance, defined by a Mann-Whitney *U* Test (*p*-value < 0.05) and fold-change > ± 1.2. A positive fold-change indicates a higher mean relative expression level in the cancer group while a negative fold-change indicates a higher mean relative expression level in the benign group.

| Antigen (Antibody Class) | $p$-value | Fold Change |
| --- | --- | --- |
| **Pro-CASP8 (IgG)** | 0.0245 | -1.31 |
| **COL1A1 (IgG)** | 0.0214 | -1.2 |
| **RHOXF2 (IgM)** | 0.0046 | 1.24 |
| **SPDEF (IgG)** | 0.0078 | 1.27 |

Examination of distributions revealed distinct patterns across TAAbs. Differences between diagnostic classes for RHOXF2 (IgM; **Fig. 2c**) and SPDEF (IgG, **Fig. 2d**) were driven by a subset of cancer donors with concentrations elevated beyond the rest of the sample distribution, while there was substantial overlap between the distribution of diagnostic classes for Pro-CASP8 (IgG; **Fig. 2e**) and COL1A1 (IgG; **Fig. 2f**). Interestingly, the IgG antibody response against both Pro-CASP8 and COL1A1 was elevated in the benign group relative to the cancer group.

Donors with elevated relative antibody expression were not conserved between different differentially expressed TAAbs (**Fig. 2g**), indicating that each TAAb provides unique information and is likely not a result of a globally elevated immune response. None of the differentially expressed biomarkers showed differences exclusively driven by one breast cancer molecular subtype, though interpretation is limited by the small subgroup sample size.

To assess potential confounding effects, we evaluated whether clinical and demographic variables influenced TAAb concentrations. Age, tumor size, tumor grade, and cancer stage were tested as potential covariates using ordinary least squares (OLS) multiple linear regression (MLR) (**SI Table 4**). Given that immune activity often declines with age, we also evaluated whether donor age influenced the relative concentrations of the identified differentially expressed TAAbs. Using the subset of TAAbs identified as significantly differentially expressed, we plotted TAAb signal intensity as a function of age (**SI Fig. 4).** No significant correlations between age and antibody response were observed (**SI Table 5**), and variance inflation factor (VIF) tests confirmed the absence of problematic multicollinearity. Cumulatively, these findings indicate that age did not account for the observed variability in TAAb distributions. Sex could not be a confounder, as all study participants were female.

Across all predictors, tumor grade was the primary variable associated with diagnostic classification, explaining the majority of variance in the model (**SI Table 6**). Notably, tumor grade showed the strongest association with individual TAAb concentrations, whereas tumor stage, tumor size, and age were non-significant once grade was included in the model. Adjusting for grade and other clinical variables, no individual TAAb remained independently associated with diagnosis (all β ∼ 0 with *p* >> 0.05). These findings indicate that TAAb signatures reflect the tumor biology captured by histological grade, rather than tumor burden (stage or size). Overall, the MLR results indicate that the observed heterogeneity in immune activity reflects intrinsic patient-specific variation rather than demographic influences or extent of disease.

Through these analyses we found that differences in relative TAAb concentration may be equally, if not more, prevalent at the individual patient level rather than as a consistent feature of diagnostic class. Additionally, we observed that some differentially expressed antibodies were elevated in donors with benign disease compared to those with cancerous disease, indicating a potential protective role of certain autoantibodies in response to neoplasms. However, the observation of patient clustering independent of diagnosis through the analysis of the heat maps and the identification of few differentially expressed biomarkers prompted evaluation of the underlying assumptions of this analysis process.

### Intra-patient normalization of immune activity reveals differentially expressed TAAbs between benign and cancer cohorts

Following the visualization of diagnosis-independent trends and the identification of few differentially expressed biomarkers in the NetI data, we investigated whether inter-individual variation in baseline immune activity masked additional diagnostic signals. To assess potential differences in total immune activity between donors, we plotted the number of IgM (**Fig. 3a**) and IgG (**Fig. 3b**) biomarkers at a given relative concentration for each donor, with distributions sorted by peak height. While most donors exhibited approximately Gaussian-like distributions with relatively consistent means, standard deviations, and distribution shapes, a subset of donors, particularly for the IgG TAAbs, exhibited substantial deviation from normality. Some donors displayed globally elevated immune responses indicative of overactive immune systems, while others exhibited narrow distributions, indicative of reduced immune activity. Additionally, five donors in the IgG panel and one donor in the IgM panel exhibited substantial variation in their antibody distribution, defined as having an individual interquartile range (IQR) exceeding the cohort-wide upper threshold (Q3 + 1.5×IQR). These differences affected the mean and standard deviation of each donor’s TAAb distribution, creating a latent variable proportional to immune activity that influenced analysis of the NetI data.

**Figure 3:**
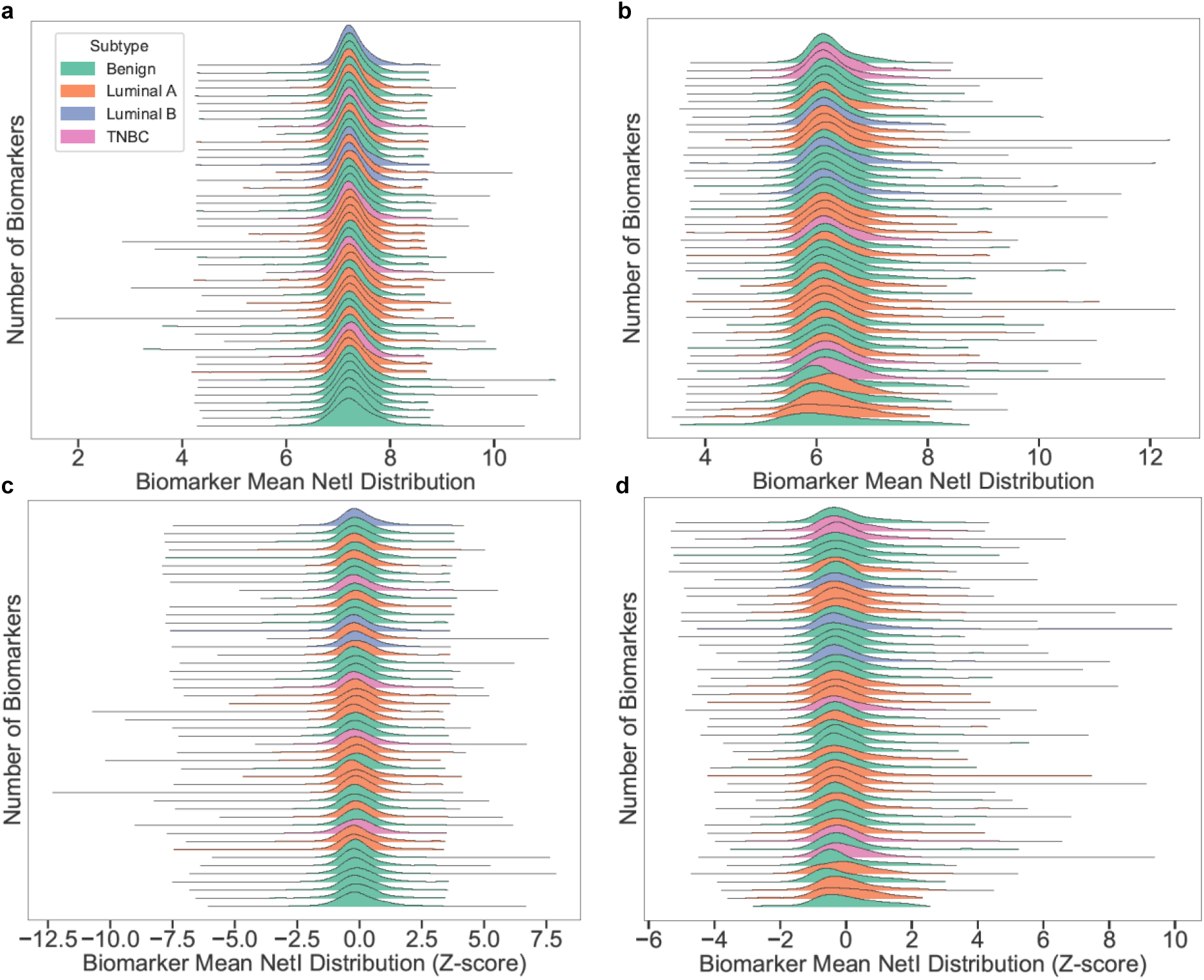
Joy plots visualize correction for variation in individual immune activity through *z*-score normalization in both the IgM and IgG panels. NetIs are plotted for both the **a)** IgM and **b)** IgG response of each donor. Each distribution represents an individual donor with color designating the diagnostic class and subtype as indicated in the legend in the top left of panel **a**. The height of the curve represents the relative number of biomarkers at a given NetI value, and the length of each distribution represents the total range of each individual’s antibody distribution. The distributions after *z*-score normalization for the **c)** IgM and **d)** IgG panels show the distributions in the same order as their corresponding NetI plot (panels **a** and **b**, respectively).

We also analyzed whether clinical factors influenced the antibody distributions, including active chemotherapy treatment at sample collection (n = 2), history of cancer in benign donors (n = 2), subsequent conversion breast cancer after sample collection in benign donors (n = 2). No obvious trends were observed in the IgM (**SI Fig. 5a**) or IgG (**SI Fig. 5b**) individual distribution shapes, indicating that the overall antibody distributions were not substantially influenced by these variables. Additionally, we found that there was no association between the mean NetI of donors’ total IgM (**SI Fig. 5c)** or IgG (**SI Fig. 5d)** TAAb distribution and disease stage. Interestingly, donors with stage III breast cancer did not have globally elevated TAAb distributions compared to donors with other disease stages or states.

To account for this variation in immune activity between patients, *z*-score normalization was applied to each donor’s TAAb distribution to calculate the “Normalized NetI” for each antibody. *z*-score normalization centers the mean of each person’s antibody distribution to zero and scales the standard deviation to one, enabling the comparison of relative antibody expression across individuals with different baseline IgM (**Fig. 3c**) and IgG (**Fig. 3d**) activities. Thus, this transformation identifies individual biomarkers that are elevated relative to each donor’s own baseline antibody distribution by expressing values as standard deviations from each individual’s mean IgM or IgG concentration, rather than comparing absolute expression titers. Moreover, *z*-score normalization prevents over-weighting individuals with unusually broad or narrow TAAb distributions from disproportionally influencing the identification of differentially expressed biomarkers between diagnostic groups. Consistent with exposure to neoantigens, we found that many donors had TAAbs that were expressed at or above 3 standard deviations above their personal mean.

After applying *z-*score normalization to account for patient specific variation in immune activity, differential expression analysis identified TAAbs elevated relative to each donor’s personal baseline immune profile (IgM: **SI Table 7**; IgG: **SI Table 8)**. A two-tailed Mann-Whitney *U* Test identified 11 IgM antibodies (**Fig. 4a**) and 14 IgG antibodies (**Fig. 4b**) that differed significantly between the benign and cancer groups *(p* < 0.05, fold-change > ± 1.2). The *p-*value and the fold-change for each of the differentially expressed biomarkers are reported in **Table 3**.

**Figure 4:**
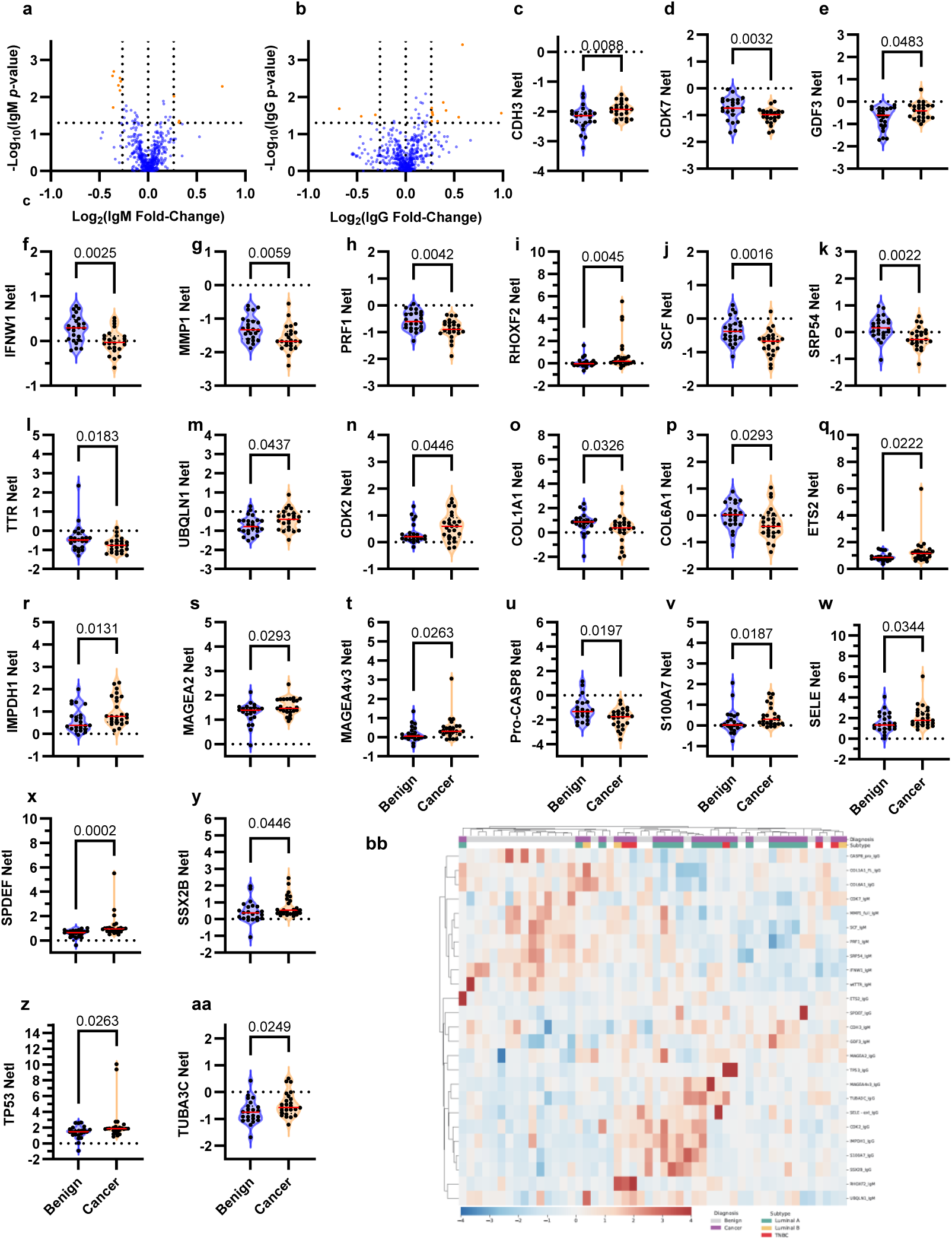
Identification of differentially expressed IgM and IgG biomarkers between benign and breast cancer donors in the Normalized NetI data set. The volcano plots show the −log_10_(*p*-value) and log_2_(fold-change) for each biomarker measured for each: **a)** the IgM response and **b)** the IgG response to 525 TAAs. Each point represents the mean of n = 4 technical replicates (n = 48 samples). The horizontal dashed line indicates statistical significance (*p*-value of 0.05). Vertical dashes indicate a fold-change > ± 1.2. Points to the left of the leftmost vertical line are biomarkers with a higher mean signal in the benign group than the cancer group, whereas points to the right of the rightmost vertical dashed line are biomarkers with higher mean signal in the cancer group. Orange points indicate biomarkers that meet both *p*-value and fold-change significance criteria. Each violin plot shows the distribution of the Normalized NetI values of each sample, distinguished by diagnostic class, for each IgM (**c-m**) and IgG (**n-aa**) biomarker identified as significant (*p* < 0.05, fold-change > ± 1.2). Each point represents the mean value of four technical replicates, or two to three if any replicates were removed for a high CV (> 20%). The *p*-values were calculated using a Mann-Whitney *U* test. The red bar shows the median. **bb)** A heat map of the relative expression of the identified significant biomarkers (rows) across all subjects (columns). Red represents high relative expression while blue represents low relative expression. Both donor and biomarker axes are clustered hierarchically using average linkage based on Pearson correlation. The diagnosis and subtype of each donor are identified in the top two rows of the plot.

**Table 3:** Summary of significant TAAbs identified by differential expression analysis of the Normalized NetI data. Displayed are the antigens to which the TAAbs are differentially expressed that meet both the *p*-value and fold-change threshold for significance, defined by a Mann-Whitney *U* Test (*p*-value < 0.05) and fold-change > ± 1.2. A positive fold-change indicates a higher mean relative expression level in the cancer group while a negative fold-change indicates a higher mean relative expression level in the benign group.

| Antigen (Antibody Class) | $p$-value | Fold Change |
| --- | --- | --- |
| Pro-CASP8 (IgG) | 0.0207 | -1.6 |
| COL1A1 (IgG) | 0.0335 | -1.37 |
| SRP54 (IgM) | 0.0027 | -1.29 |
| SCF (IgM) | 0.0021 | -1.28 |
| TTR (IgM) | 0.0193 | -1.28 |
| PRF1 (IgM) | 0.0049 | -1.24 |
| COL6A1 (IgG) | 0.0302 | -1.24 |
| CDK7 (IgM) | 0.0038 | -1.22 |
| IFNW1 (IgM) | 0.0031 | -1.22 |
| MMP1 (IgM) | 0.0067 | -1.22 |
| CDH3 (IgM) | 0.0097 | 1.2 |
| CDK2 (IgG) | 0.0453 | 1.2 |
| S100A (IgG) | 0.0197 | 1.2 |
| MAGEA2 (IgG) | 0.0302 | 1.21 |
| MAGEA4v3 (IgG) | 0.0272 | 1.21 |
| TUBA3C (IgG) | 0.0258 | 1.21 |
| GDF3 (IgM) | 0.0489 | 1.24 |
| UBQLN1 (IgM) | 0.0444 | 1.25 |
| SSX2B (IgG) | 0.0453 | 1.26 |
| IMPDH1 (IgG) | 0.0141 | 1.31 |
| ETS2 (IgG) | 0.0232 | 1.33 |
| SELE (IgG) | 0.0353 | 1.46 |
| SPDEF (IgG) | 0.0004 | 1.5 |
| RHOXF2 (IgM) | 0.0052 | 1.7 |
| TP53 (IgG) | 0.0272 | 1.98 |

The significant IgM antibodies were against Cadherin 3 (CDH3), Cyclin-Dependent Kinase 7 (CDK7), Growth Differentiation Factor 3 (GDF3), Interferon Omega 1 (IFNW1), Matrix metalloproteinase-1 (MMP1), Perforin 1 (PRF1), RHOXF2, Mast cell growth factor (SCF), Signal Recognition Particle 54 (SRP54), Transthyretin (TTR), and Ubiquilin-1 (UBQLN1) (**Fig. 4c-m**). The significant IgG antibodies were against Cyclin-Dependent Kinase 2 (CDK2), COL1A1, Collagen Type VI Alpha 1 Chain (COL6A1), ETS Proto-Oncogene 2 (ETS2), Inosine Monophosphate Dehydrogenase 1 (IMPDH1), MAGE Family Member 2 (MAGEA2), MAGE Family Member 4 (MAGEA4v3), Pro-CASP8, S100 Calcium Binding Protein A7 (S100A7), E-selectin (SELE), SPDEF, SSX Family Member 2 (SSX2B), Tumor Protein 53 (TP53), and Tubulin alpha-3C chain (TUBA3C) (**Fig. 4n-aa**). The biomarkers that were differentially expressed in the NetI data were conserved in analysis of the Normalized NetI analysis, and 21 additional biomarkers met the significance criteria with this normalization approach.

Examination of individual antibody distributions revealed heterogenous expression patterns. Similar to the analysis of the NetI data, we observed that many of the differentially expressed antibodies were not globally elevated as a function of diagnostic class but rather were highly expressed in specific patients, suggesting patient-specific immune activation. Some antibodies have differences in fold-change driven by shifts in the total distributions of the diagnostic classes, including CDK7, IFNW1, and PRF1 in the IgM group (**Fig. 4d, f, and h**) and CDK2, IMPDH1, and SSX2B in the IgG group (**Fig. 4n, r, and y**). Others have differences in fold-change driven primarily by individual donors with high expression levels, such as RHOXF2 and TTR (IgM; **Fig. 4i** and **l**) and ETS2, SPDEF, and TP53 (IgG; **Fig. 4q, x, and z**). For example, the IgG response to CDK2 (**Fig. 4n**) was tightly clustered among benign donors, with only a small subset (n = 5) exhibiting elevated antibody levels, whereas cancer samples displayed a broader distribution with numerous donors showing comparatively higher titers. This pattern of patient-specific immune activation was consistent with observations from the NetI data. Notably, many differentially expressed biomarkers were elevated in the benign group compared to the cancer group (IgM n = 7; IgG n = 3), supporting the idea that some TAAbs may play a protective role against the development of malignancies rather than an oncogenic role. The donors with elevated high relative expression levels were not conserved between different TAAbs, and none of the differentially expressed TAAbs have differences exclusively driven by one molecular subtype (**Fig. 4bb**).

Pathway analysis of the differentially expressed TAAbs identified through analysis of the Normalized NetI data revealed functional clustering centered on TP53 (**SI Fig. 6**), consistent with the widely accepted role of TP53 pathway dysregulation in cancer development and progression^48^. Network analysis using STRING-DB demonstrated that 16 of the 25 differentially expressed TAAbs map to TP53-associated signaling pathways, including DNA damage response, cell cycle regulation, and apoptosis control^49^. While each individual displayed a unique antibody signature and no single TAAb was able to differentiate benign donors from cancer donors, the functions of antigens targeted by differentially expressed TAAbs converge on the TP53 signaling pathway. Notably, antibodies against three extracellular matrix and remodeling proteins, MMP1, COL1A1, and COL6A1, are upregulated in the benign class. Given that these proteins are implicated in tumor metastasis and cell invasion, their elevated titer in donors with benign disease supports the idea that an immune response against certain tumorigenic antigens may be preventative against disease progression to cancer.

Although the specific differentially expressed antibodies differed between isotypes, pathway analysis revealed conserved biomarker families and functional pathways between IgM and IgG responses. For example, TAAbs against cyclin-dependent kinases were differentially expressed in both isotypes (IgM: CDK7; IgG: CDK2), as were TAAbs against extracellular matrix remodeling proteins (IgM: MMP1; IgG: COL1A1). These isotype-specific differences between differentially expressed TAAbs may reflect the downregulation of the initial IgM antibodies prior to the development of an IgG response rather than a total lack of conservation of antibody isotypes. This conservation of responses to related TAA families across isotypes suggests that evaluating multiple antibody isotypes may provide a more complete picture of anti-tumor immunity and better reflect the diagnostic status of an individual. Additionally, IgM antibodies were primarily elevated in the benign diagnostic class (7 of 11 total differentially expressed antibodies), which may indicate that donors in this class are able to clear elevated TAAs with the initial IgM antibody response more effectively than donors in the cancer class.

Parallel to the analysis of the NetI data, we evaluated whether clinical and demographic variables influenced the differentially expressed TAAbs identified in the normalized dataset. Age, tumor size, tumor grade, and tumor stage were evaluated as potential confounding variables using OLS MLR (**SI Table 9**). No significant correlations were observed between age and any of the differentially expressed TAAbs (**SI Fig. 7, SI Table 10**), consistent with the NetI analysis. VIF testing revealed modest multicollinearity with tumor grade exhibiting the expected correlation with stage and size. Two antibodies (S100A7 IgG and IMPDH1 IgG) exhibited VIF > 10, suggesting redundancy with other immune markers; however, excluding either of these high-VIF variables did not affect the model outcomes.

MLR analysis of the Normalized NetI dataset demonstrated that the 25 differentially expressed TAAbs collectively accounted for 97.7% of the diagnostic variance (R^2^, F-statistic *p* << 0.001). Tumor grade remained the most dominant correlate with diagnosis (*p* << 0.001), while age, tumor size, and tumor stage were not significant when accounting for other covariates. Notably, a subset of TAAbs had independent associations with diagnosis after controlling for clinical covariates. Five antibodies were positively correlated with the cancer diagnostic class, ranked by the model coefficient: anti- MAGEA4 IgG, CDK2 IgG, MMP1 IgM, CDH3 IgM, and UBQLN1 IgM (all *p* < 0.05). Conversely, four antibodies showed significant negative correlations with diagnosis: anti- S100A7 IgG, SCF IgM, CDK7 IgM, and RHOXF2 IgM (all *p* < 0.05), with TP53 IgG approaching significance (*p* = 0.055). It should be noted that MLR coefficients represent associations after controlling for all covariates and often diverge from univariate expression patterns. For example, anti-MMP1 IgM has a positive MLR coefficient despite showing higher absolute expression in benign samples, while anti-S100A7 IgG and anti-RHOXF2 IgM have negative MLR coefficients but are overexpressed in cancer samples. In contrast, other antibodies have coefficients aligned with the expected diagnostic class. These discordant patterns highlight that MLR captures complex multivariate relationships rather than simple univariate associations. These autoantibody signatures likely reflect the immune system’s response to differential antigen expression between benign and malignant tissue. Elevated autoantibodies may indicate either successful immune surveillance against their cognate antigens or, alternatively, may arise in response to antigen overexpression associated with tumor development. The biological significance of these immune responses—whether protective, reactive, or pathogenic—requires further investigation with antigen expression data and functional studies. In contrast to the NetI analysis, where no TAAbs had independent associations after adjusting for grade, these associations persisted after adjusting for grade and other clinical variables, demonstrating that intra-patient normalization reveals TAAb signatures reflecting tumor biology beyond histological grade alone (**SI Table 11)**. These differentially expressed biomarkers form the basis for subsequent evaluation of their distribution patterns and classification performance.

### Benign breast disease is associated with heightened immune activation relative to breast cancer

To determine whether donors with breast cancer had a heightened cancer-related immune profile relative to donors with benign disease, we quantified the magnitude of immune activation in each diagnostic class (IgM: **Fig. 5a**; IgG: **Fig 5b**). For each donor, the number of “high-titer” antibodies was calculated in both the IgM and IgG panels. In this study, a high-titer antibody was defined as an antibody with a Normalized NetI greater than three standard deviations above the cohort mean. This conservative, distribution-based cutoff was chosen for its simplicity and reproducibility across a large antibody panel, in contrast to antigen-specific ROC-derived thresholds that require individual optimization. As with biomarker identification, conducting this analysis following normalization prevents bias from patients with a globally elevated immune response or with especially wide antibody distributions.

**Figure 5:**
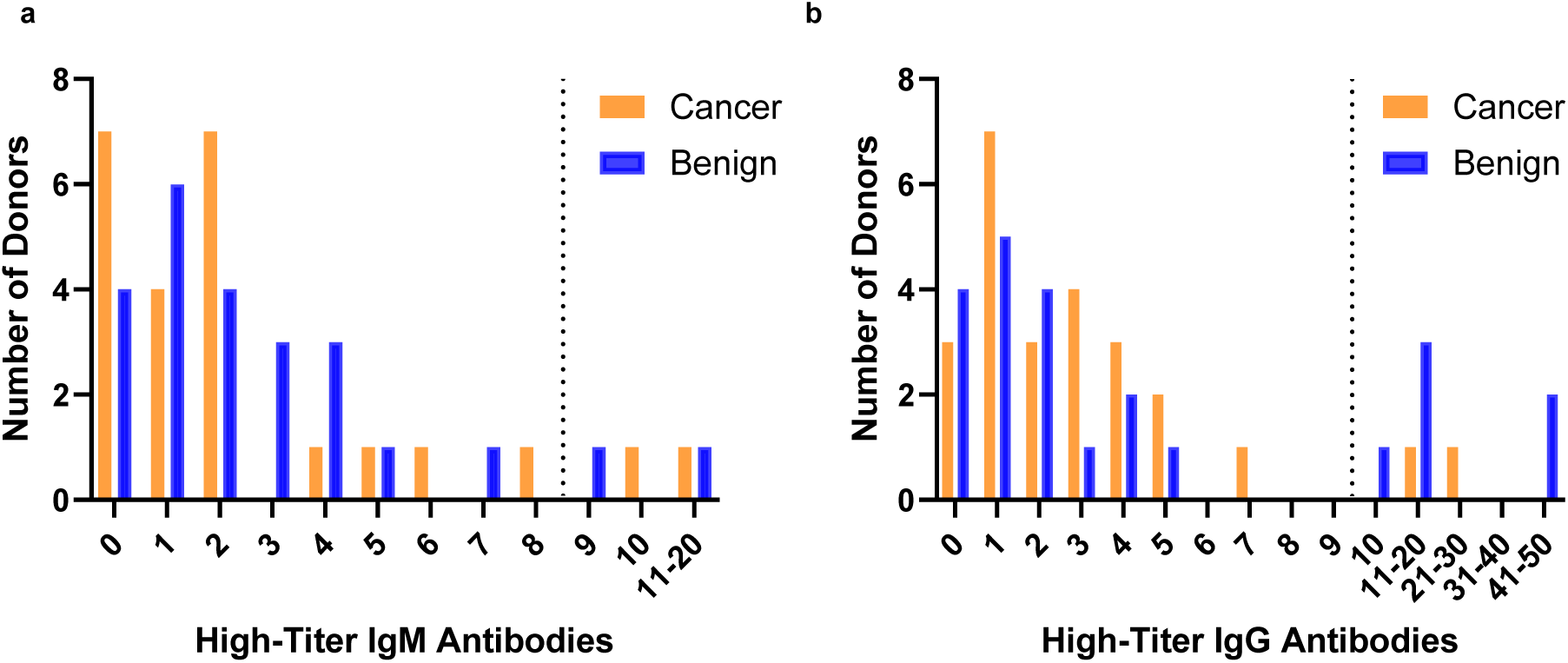
Benign donors exhibit elevated IgG responses when evaluated for high-titer antibodies. Histogram plots of the number of high-titer antibodies per donor in the **a)** IgM and **b)** IgG panels following *z*-score normalization. The vertical dashed line on each plot indicates the cohort-wide upper threshold (Q3 + 1.5×IQR), defining donors with an elevated immune response.

Interestingly, the benign class has a higher prevalence of donors with elevated numbers of high-titer antibodies in the IgG (**Fig. 5b**) panel than the cancer class. In the IgM panel, the distributions are similar between classes, with both groups forming narrow right-skewed distributions (Benign: median = 2, IQR 0-2.5; Cancer: median = 2, IQR 1-4). Two donors in each diagnostic class have elevated IgM profiles, characterized by a number of high-titer antibodies exceeding the cohort-wide upper threshold (Q3 + 1.5×IQR). However, in the IgG panel, the cancer group has a narrow Gaussian-like distribution (median = 2, IQR 1-4), with two donors having an elevated IgG profile. Of these two outliers, one was the only donor receiving chemotherapy treatment at the time of sample collection, which may have affected the donor’s immune activity. The benign group demonstrated a wider range of high-titer IgG antibodies (median = 2, IQR 1-7.5) with six donors expressing an elevated IgG profile. There was no correlation observed between the number of high-titer IgM and IgG antibodies at the donor level (R^2^ = 0.006), indicating that everyone exhibits a unique isotype response.

Together, these findings demonstrate that benign breast disease is often accompanied by an activated humoral immune response, with benign donors exhibiting a greater number of high-titer IgG TAAbs than cancer donors. Interestingly, this pattern emerged following within-patient normalization, indicating that heightened self-relative immune activity may characterize certain benign or pre-malignant states, whereas malignancy may be associated with comparatively dysregulated or suppressed antibody responses. In this context, strong antibody responses against tumor-associated antigens may serve a protective role.

Network analysis using STRING-DB was performed on the high-titer antibodies of individual donors to identify whether there were trends in immune profiles between donors. Interestingly, there did not appear to be a single dominant pathway shared across donors; instead, each donor exhibited a distinct immune pattern. For example, in the analysis of donors’ high-titer IgG antibodies, one benign donor showed broad enrichment for cytokine- and chemokine-associated signaling, with elevated antibodies targeting IL6ST, multiple interleukins (*e.g.*, IL12A, IL13, IL23A), and chemokines (*e.g.*, CCL25, CXCL13), consistent with a predominantly inflammatory and JAK–STAT–driven immune profile (**SI Fig. 8a**). In contrast, a second benign donor displayed a different TAA recognition pattern, with high-titer IgG antibodies recognizing proteins involved in proliferative signaling (*e.g.*, AKT1, MAPK1/3), apoptosis regulation (*e.g.*, BIRC5, CASP3), and numerous cancer-testis antigens (*e.g.*, MAGE family members, PAGE family members, GAGE4). This profile was instead dominated by pathways related to MAPK signaling, cell-cycle control, and tumor-associated antigen expression (**SI Fig. 8b**). These two donors were selected for in-depth analysis as they had the highest number of high-titer IgG antibodies. Moreover, the antigens targeted by high-titer antibodies differ within diagnostic classes, reflecting the heterogeneity of genesis and progression in both malignant and benign breast disease. These findings suggest that high-titer antibody profiles do not share a single biological pathway across donors, but rather, each donor’s serologic signature reflects a personalized combination of inflammatory, proliferative, or tumor-associated processes. Additionally, the higher frequency of benign donors with robust immune responses to pathways commonly implicated in oncogenesis and disease progression supports the well-studied hypothesis of immune surveillance and evasion, as these individuals are better able to mount an effective immune response against early cancer cells.

### Logistic regression classification demonstrates diagnostic potential of a combined IgM and IgG TAAb signature

We first applied unsupervised principal component analysis (PCA) to the Normalized NetI datasets to visualize global patterns of TAAb expression and assess whether benign and cancer samples exhibited distinct clustering. PCA was applied to all 525 TAAb antigens in the IgM (**Fig 6a**), IgG (**Fig. 6b**), and combined IgM and IgG panels (**Fig. 6c**). No distinct clustering of donors by diagnostic class was observed along the first two principal components for any dataset, indicating that the antibody expression patterns were broadly similar across the groups. Cancer subtypes likewise lacked distinct patterns of immune reactivity, with luminal A, luminal B, and triple-negative breast cancers dispersed without obvious clustering (orange, blue, and pink, respectively). Scree plots indicated that approximately 30% of the total IgM variance **(SI Fig. 9a**), 40% of the total IgG variance (**SI Fig. 9b**), and 27% of the total combined IgM and IgG variance (**SI Fig. 9c**) were captured by the first two components, reflective of the diversity and quantity of antibodies analyzed in this study. These findings suggest that the total TAA-related immune response lacks a cancer-specific structure, is highly individualized rather than class driven, and reinforces the need to narrow to differentially expressed antibodies to distinguish diagnostic classes and enable classifier development.

**Figure 6:**
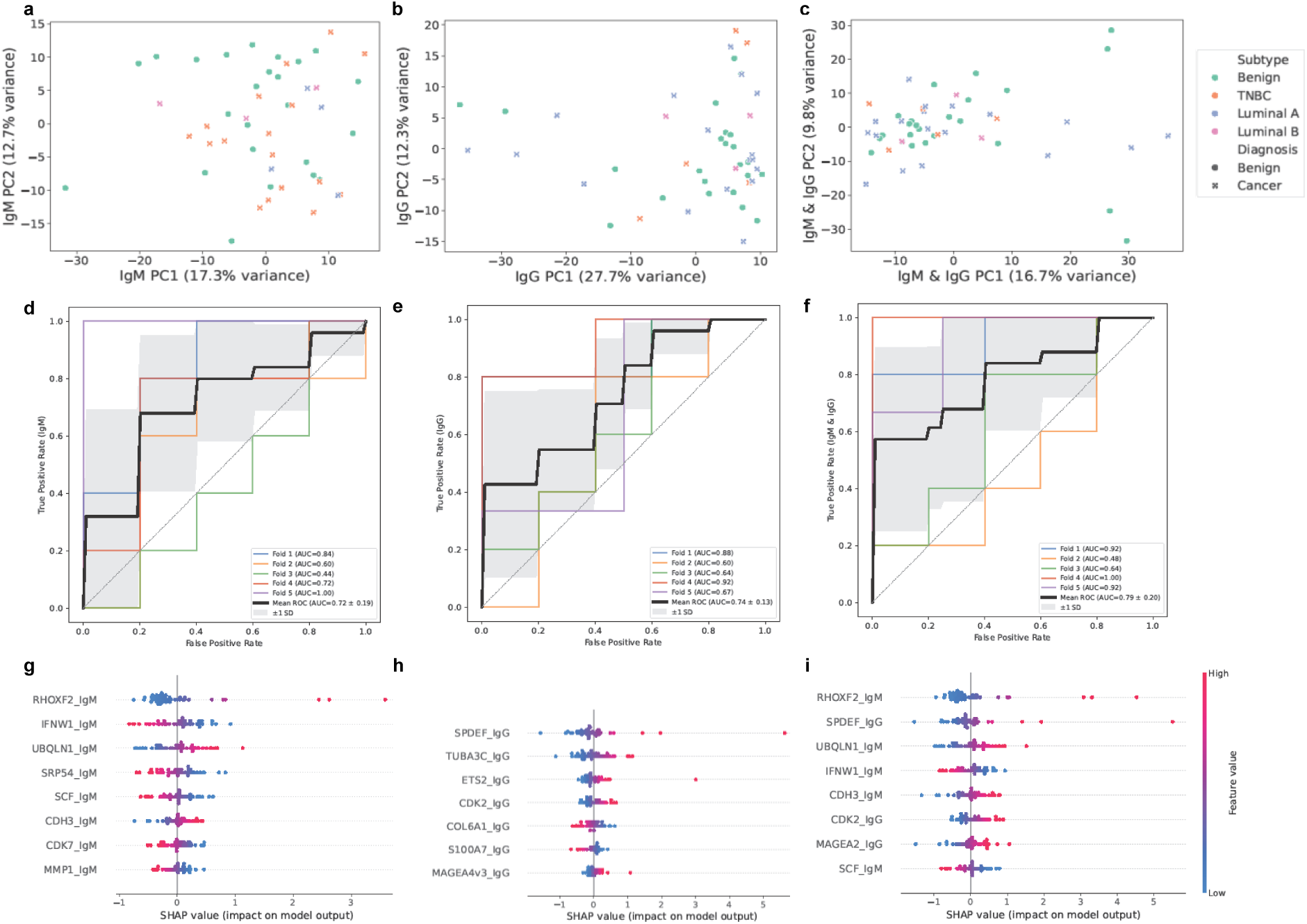
Combined IgM and IgG TAAb panel improves classification of benign from malignant disease. PCA plots of Normalized NetI for all 525 TAAb antigens for **a)** IgM, **b)** IgG, and **c)** combined IgM and IgG TAAb panels show partial separation of benign samples (green circles) and cancer samples (crosses, colored by subtype: orange, luminal A; blue, luminal B; pink, triple negative). ROC curves for logistic regression classifiers built from the top-performing biomarkers selected via nested recursive feature elimination for the **d)** IgM, **e)** IgG, and **f)** combined IgM and IgG panels. Each fold of the five-fold cross-validation is shown by a different color; the bold black line represents the mean ROC across folds. Shaded gray area indicates the 95% confidence interval. SHAP beeswarm plots illustrate feature contributions to model predictions for **g)** IgM, **h)** IgG, and **i)** combined IgM and IgG classifiers. Colors indicate feature value (red, high relative Normalized NetI; blue, low relative Normalized NetI). Features with positive SHAP values (right of the vertical line) contribute to cancer prediction, while negative SHAP values (left of the vertical line) contribute to benign predictions.

Limiting further analysis to the differentially expressed antibodies, we evaluated whether combinations of the most significant TAAbs could distinguish benign from cancer samples using supervised multivariate classification. To minimize overfitting due to the limited sample size and large number of antibodies analyzed, only the TAAbs identified as differentially expressed and the absolute number of high-titer antibodies were included in the building of classifiers (28 possible features; n = 11 IgM; n = 14 IgG, age, and the number of high-titer IgM or IgG antibodies). Clinical covariates except age were excluded as they would not be available at the time of a mammogram or biopsy.

Logistic regression classifiers were trained on the selected features for the IgM, IgG, and combined panels using nested five-fold cross-validation with stratified sampling. Recursive feature elimination (RFE) was employed within the inner loop to identify the top-performing features that maximized AUC-ROC (**Fig. 6d-f)**. Feature selection and feature importance plots (**SI Figure 10a-f**) guided the selection of the optimal number of features for each classifier. Through this analysis, 8 features were selected for the IgM classifier (anti- RHOXF2, IFNW1, UBQLN1, SRP54, SCF, CDH3, CDK7, and MMP1), 8 features were selected for the IgG classifier (anti-SPDEF, TUBA3C, ETS2, CDK2, COL6A1, S100A7, and MAGEA4), and 8 features were selected for the combined classifier (IgM: anti- RHOXF2, UBQLN1, IFNW1, CDH3, SCF; IgG: anti-SPDEF, CDK2, MAGEA2).

The resulting models demonstrated promising discriminatory performance despite the limited sample size. Classifiers built from the IgM panel (**Fig. 6d**) and IgG panel (**Fig. 6e**) each achieved moderate separation between diagnostic groups (mean AUC-ROC = 0.78 ± 0.17 and 0.70 ± 0.13, respectively, averaged across 10 repetitions of five-fold cross-validation), whereas a combined model incorporating both IgM and IgG biomarkers yielded the best overall performance (mean AUC-ROC = 0.83 ± 0.14; **Fig. 6f**). The ROC curves illustrated represent the mean performance across a single five-fold cross-validation. These findings indicate that an integrated, self-relative IgM and IgG TAAb signature provides diagnostic value and may serve as a foundation for development of serological classifiers of early-stage breast cancer detection.

SHAP analysis was performed on the final trained classifiers using the LinearExplainer to interpret feature contributions and visualize the directionality of each TAAb in relation to diagnostic class. In the IgM panel (Fig. 6g), five of the eight selected TAAbs showed elevated expression in the benign group with varying levels of feature importance (anti- IFNW1, SRP54, SCF, CDK7, and MMP1), supporting that certain IgM responses may be associated with benign conditions. Conversely, elevated anti- RHOXF2, UBQLN1, and CDH3 were associated with cancer classification. In the IgG panel (Fig. 6h), high levels of anti- COL6A1 and S100A7 were associated with benign classification, while the remaining IgG features (anti- SPDEF, TUBA3C, ETS2, CDK2, and MAGEA4) showed the opposite pattern, with higher expression corresponding to increased cancer probability. This contrasts with the predominantly benign-associated IgM signature, suggesting distinct functional roles for the two antibody isotypes. In the combined classifier (Fig. 6i), elevated anti- RHOXF2, SPDEF, UBQLN1, CDH3, CDK2, and MAGEA2 were associated with cancer classification, while high anti- IFNW1 and SCF levels were linked to benign status. The bidirectional nature of these associations with some TAAbs elevated in benign samples and others in cancer, demonstrates that diagnostic information is encoded not only in the presence of the TAAb, but also in the direction of their relative expression patterns. Notably, 5 of the 8 final classifier biomarkers showed direct, secondary, or tertiary interactions with anti-TP53, suggesting that, while TAA recognition varies widely between individuals, recognition of stress-response and tumor-suppressor pathway components are functional pathways that are commonly able to distinguish disease states. These findings highlight the complexity of humoral immune responses in breast disease and underscore the value of multivariate classification approaches that capture both protective and disease-associated antibody signatures.

## Discussion

In this study, we analyzed serum IgM and IgG responses to 525 TAAs in 50 donors (26 with predominantly early-stage breast cancer and 24 with benign breast disease) to identify antibody signatures capable of distinguishing malignant from benign disease. Through *z*-score normalization of each donor’s personal antibody distribution, we accounted for inter-individual variation in immune activity, allowing identification of antibodies that were elevated relative to each donor’s baseline immune profile. This self-normalization approach uncovered 25 differentially expressed TAAbs and enabled the development of a predictive model integrating eight antibodies that achieved an AUC-ROC of 0.83, indicating measurable diagnostic potential despite the limited cohort size. Critically, this comparison between early-stage cancer and benign disease addresses a diagnostic challenge that has been understudied relative to the more common cancer-versus-healthy comparisons in the literature.

While no single TAAb provided sufficient discriminatory power, combining multiple features improved diagnostic accuracy and captured the biological complexity of breast cancer. We observed two distinct immunological patterns: 1) the total number of high-titer autoantibodies across the panel and 2) the expression levels of specific individual autoantibodies. Regarding the first pattern, we observed benign donors exhibited a significantly higher frequency of high-titer antibodies for the IgG isotype compared to cancer patients. This suggests that individuals with benign breast disease may mount a more effective immune response to nascent tumor antigens, representing an innate protective mechanism that prevents malignant transformation. However, the total number of high-titer antibodies was not directly diagnostic in our classifier. Regarding the second immunological pattern of autoantibody specific differences, elevated concentrations of specific IgM antibodies were predominantly associated with benign disease, whereas increased levels of specific IgG antibodies more often predicted malignancy. These opposing patterns may reflect distinct functional roles—IgM-mediated early surveillance versus IgG responses that either fail to contain or inadvertently promote tumor progression, or increased concentrations that arise due to sustained antigen exposure from an established malignancy. While the total high-titer antibody count did not improve classifier performance, individuals with benign disease who exhibit heightened IgG antibody responses may have enhanced capacity to prevent progression to malignancy. This observation warrants investigation of whether high-titer antibody profiles predict cancer-free survival.

Pathway analysis identified functional links of antigens targeted by differentially expressed TAAbs enriched in the TP53 signaling pathway, consistent with the widely accepted role of cell cycle, DNA repair, and apoptotic dysregulation in oncogenesis. Moreover, since TP53 is a cell-cycle regulatory protein with tumor-suppressive properties, the upregulation of TP53 TAAbs restricted to two donors in the cancer group of this study supports the hypothesis that high expression of antibodies against tumor-suppressive proteins may play an oncogenic role^50^. This finding, specifically, is supported by other studies investigating the role of TAAbs in breast cancer development and progression^14,17^. Conversely, it is possible that TAAbs repress certain proteins that play a role in disease progression, such as CDK7, which may help to maintain a benign disease state. Whether the identified differentially expressed antibody responses in this study reflect attempted immune containment of early malignant changes or instead indicate immune dysregulation that permits tumor escape remains unclear and warrants mechanistic investigation in a larger study. Antibodies against proteins involved in oncogenic processes, including anti-CDH3^51^ and anti-MMP1^52^ (cell adhesion and extracellular matrix remodeling), and antibodies targeting MAGEA family members^29^ (cancer-testis antigens) were amongst those identified in this analysis. Notably, anti-CDH3 IgM and anti-MMP1 IgM were among the early-response antibodies identified, which suggests that immune recognition of these targets may provide early signals of malignant transformation. Whether repression of these antibody targets contributes to tumor development or instead reflects an early immune attempt at containment remains to be clarified but may relate to the role of the specific antigen or the response to an individual’s distinct disease pathway.

This cohort offers a rare, well-controlled comparison of early-stage breast cancer and benign breast disease. The cancer group consisted predominantly of stage I (n = 10) and stage II (n = 4) patients, with smaller representation of stage III or IV (n = 4) or unknown stage (n = 7), enabling a focus on early disease where diagnostic ambiguity is greatest and serological biomarkers could have maximal clinical impact. Critically, all samples were collected and processed using identical protocols by the same study staff, minimizing pre-analytical variability that may have confounded prior TAAb studies. Consequently, this data set provides a controlled platform for exploring immune-based differentiation of breast disease states, addressing a longstanding limitation of prior TAAb studies that compared cancer to healthy controls rather than benign disease, despite the clinical reality that distinguishing malignant from benign lesions represents the primary diagnostic challenge following abnormal imaging findings.

A key methodological insight from this work is the value of self-normalization in antibody-based diagnostics. Traditional population-level reference intervals assume a uniform biological baseline, which is rarely the case for any biomarker, and especially so for immune-responses shaped by individual exposure and history. The *z*-score normalization across each donor’s total antibody distribution accounts for patients with both globally hyperactive and hypoactive immune states, enabling per-patient interpretation of relative antibody elevation. This principle may inform future personalized diagnostics in which reference ranges are individualized rather than population derived. Because the *z*-score normalization is inherently self-referential, it could also enable longitudinal, patient-specific monitoring, allowing individuals to serve as their own baseline over time. Such an approach could detect immune shifts associated with disease recurrence or progression prior to clinical detection and may identify changes that precede malignant transformation in high-risk individuals.

Despite these promising findings, this study has several limitations. The small sample size (n = 50) constrains statistical power and increases false discovery risk, particularly given the large screening panel of 525 antigens per isotype. While this breadth was essential for discovery and enabled the self-normalization approach, clinical translation will require down-selection to a practical subset compatible with lower-multiplexing, cost-effective technologies. This reduction will require identification of stable host reference antibodies that perform consistently across diverse populations or necessitate development of alternative normalization strategies that maintain the self-referential principle with fewer markers. Additionally, the cohort lacks HER2-enriched (ER-/PR-negative) tumors, limiting assessment of subtype-specific immune signatures across the full spectrum of molecular subtypes. Moreover, while the specificity of this screening panel to cancer-related targets was well-suited for the identification of tumor-related antibodies, no other cancers were analyzed to assess the specificity of the identified antibody targets to breast cancer. Validation in larger, independent cohorts will be essential to confirm reproducibility and assess generalizability.

Future studies should expand in five key directions. First, longitudinal sampling will be critical to determine whether heightened humoral responses, particularly IgM-dominant signatures, correlate with protection against malignant transformation or improved prognosis. Multi-year longitudinal studies could reveal whether persistent high-titer antibody responses correlate with cancer-free survival and clarify whether TAAb responses represent effective immune surveillance or simply reflect chronic antigenic exposure from benign lesions. Second, further analysis of antibody isotype subclasses (*e.g.*, IgG_1_, IgG_2_, IgG_3,_ IgG_4_) and additional antibody isotypes, such as IgA, should be conducted to determine if there is a more diagnosis-specific immune response. Third, the current platform quantifies total antibody concentrations but cannot resolve functional effector states. Post-translational modifications such as Fc sialylation can convert antibodies from pro-inflammatory to anti-inflammatory, potentially creating a tumor-protective environment rather than mediating clearance. Future studies should assess antibody quantity, antigen specificity, and include glycosylation profiles to determine whether identified TAAbs are immunologically active or functionally inert. Fourth, applying the same self-normalization technique to datasets including healthy controls may reveal whether TAAb elevation can distinguish normal immune states from benign disease-associated activation, clarifying if the elevated antibody responses in benign patients emerge in conjunction with tissue changes or reflect pre-existing immune capacity. Finally, integration of TAAb profiles with antigen quantification, radiological image findings, additional biomarkers, and additional health record information could enhance diagnostic precision and enable personalized screening. TAAbs represent one class of measurable biomarkers that also includes proteins and nucleic acid sequences. Integrated classifiers could identify high-risk individuals who would benefit from intensified screening protocols, while sparing low-risk patients from unnecessary interventions, associated healthcare costs, and callback anxiety.

## Conclusions

In summary, this study demonstrates that personalized, self-normalized analysis of tumor-associated autoantibodies can reveal disease-relevant immune signatures that are obscured in population-level comparisons. The unexpected finding that benign donors exhibit broadly elevated antibody responses, yet isotype-specific directional associations exist at the individual antigen level, highlights the complexity of the immune response to neoplastic growth. By combining immune profiling with individualized normalization and multivariate classification, these results demonstrate the feasibility of developing serological, patient-normalized classifiers for early breast cancer detection. The principle of self-normalization demonstrated here may extend beyond TAAb analysis to other high-dimensional biomarker platforms where inter-individual baseline variation confounds population-level interpretations. With validation in larger cohorts and integration with complementary diagnostic modalities, this approach could contribute to personalized screening strategies that account for individual immune variability, ultimately improving early detection accuracy and reducing overdiagnosis in breast disease.

## Data Availability

All data produced in the present study are available upon reasonable request to the authors. Upon publication, all data generated or analyzed during this study will be included in the published article and its supplementary information files.

## List of Abbreviations

AUC-ROC: Area under the curve of the receiver operating characteristic
ER: Estrogen receptor
IQR: Interquartile range
MLR: Multiple linear regression
NetI: Net intensity
OLS: Ordinary least squares
PCA: Principal Component Analysis
PR: Progesterone receptor
PSA: Poly-specific antibodies
RFE: Recursive feature elimination
RFECV: Recursive feature elimination with cross-validation
RFI: Relative fluorescent intensity
SHAP: SHapley Additive exPlanations
TAA: Tumor-associated antigen
TAAb: Tumor-associated autoantibody
TNBC: Triple-negative breast cancer
TNM: Tumor, node, metastasis
VIF: Variance-inflation factor

## Declarations

### Ethics Approval and Consent to Participate

Written informed consent was obtained from all donors in accordance with Mass General Brigham Institutional Review Board Protocol # 2022P000451.

### Availability of Data and Materials

All data generated or analyzed during this study are included in this published article and its supplementary information files. The underlying code and example datasets for this study is available in a publicly available GitHub repository and can be accessed via this link: https://github.com/Walt-Lab/XXX (URL to be made public upon publication).

### Authors’ Contributions

K.A.L. and J.C.R designed the experiments, analyzed results, and prepared the manuscript. D.R.W. supervised the research and edited the manuscript. All authors reviewed and approved the final manuscript.

## Acknowledgements

Authors thank Deborah A Dillon, MD, Cory R Weiss, MD, and Sarah Green for their invaluable assistance in collecting the samples used in this study; and Chih-Ping Mao, MD, PhD for insightful discussions.

