## Supplemental Information for "Personalized Autoantibody Profiling Distinguishes Early-stage Breast Cancer from Benign Disease"

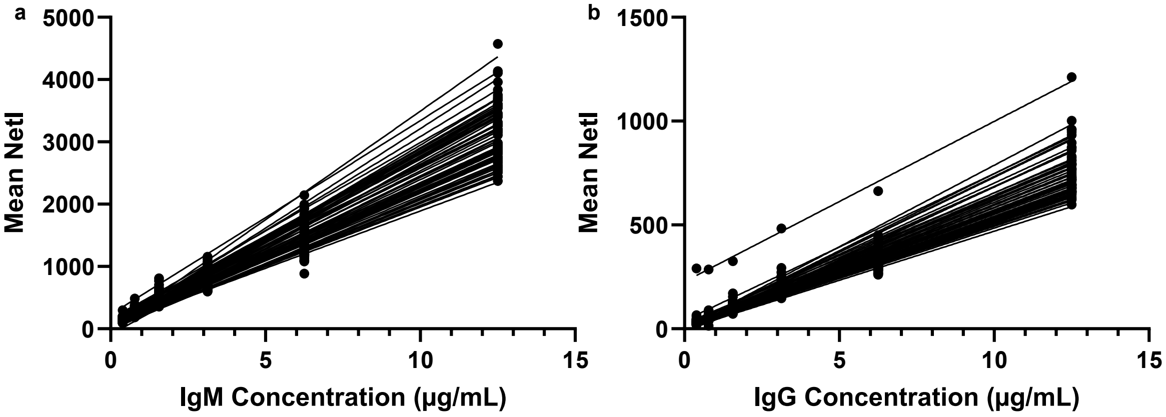

**SI Figure 1: Serial dilutions of IgG and IgM validate linearity of sample dilution and performance of secondary antibody.** Known concentrations of both **a)** IgM and **b)** IgG were titrated in a 2-fold serial dilution starting from 12.5 µg/mL on each array to validate both the linearity of the dilution as well as the performance of the secondary antibody to detect its target (n = 58).

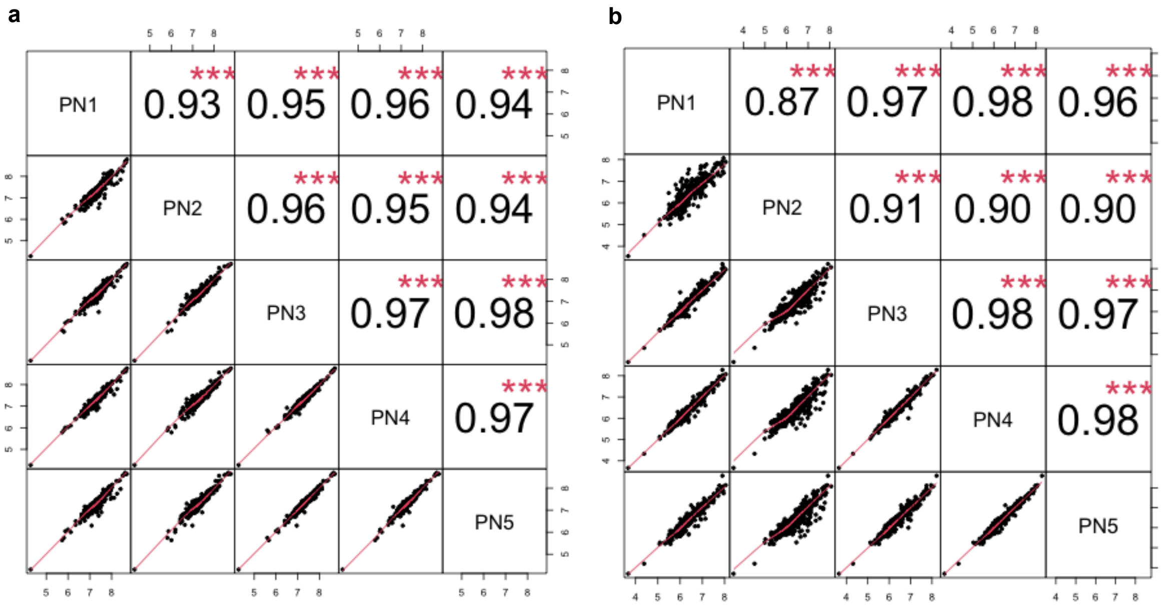

**SI Figure 2: Correlation of pooled normal biological replicates following array-Loess normalization validates normalization procedure.** For each pairwise combination of the five pooled normal samples in the **a)** IgM and **b)** IgG panel, the NetI for all antibodies were plotted against each other and the Pearson correlation coefficient (r) was calculated. Each plot is labeled according to the samples represented in the corresponding row and column, with sample identifiers displayed along the diagonal.

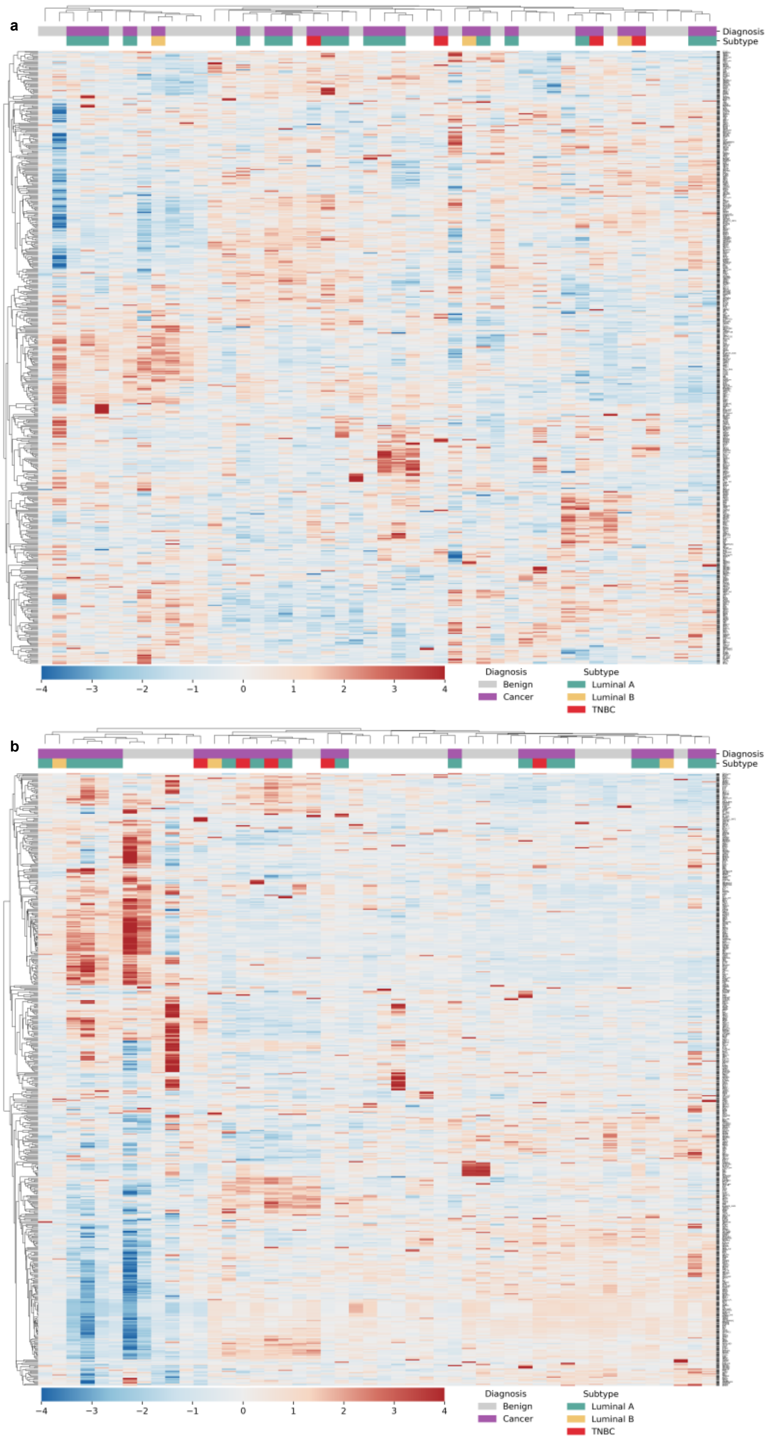

**SI Figure 3: Heatmaps show limited diagnosis- or subtype-associated trends in NetI data.** Heatmaps show the relative expression of all 525 **a)** IgM and **b)** IgG antibodies (rows) across all donors (columns). Red indicates high relative expression, while blue indicates low relative expression. Both donor and biomarker axes are clustered hierarchically using average linkage based on Pearson correlation. Donor diagnosis and molecular subtype are annotated in the top two rows of each plot.

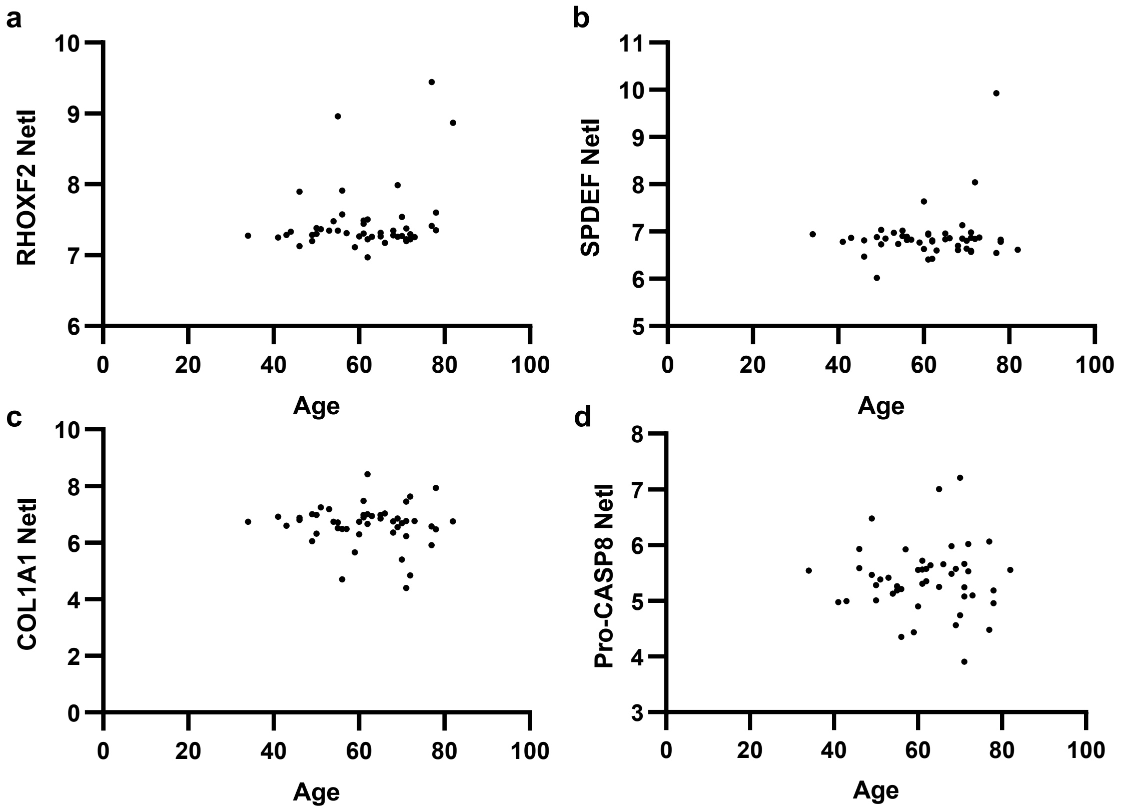

**SI Figure 4: No correlation is observed between donor age and antibody expression levels for differentially expressed antibodies identified from analysis of the NetI data.** Each point represents an individual donor, with NetI values for each differentially expressed IgM (**a**) or IgG (**b-d**) antibody NetI plotted against donor age. The NetI values represent the average of four technical replicates (or two to three if any replicates were removed for a CV > 20%).

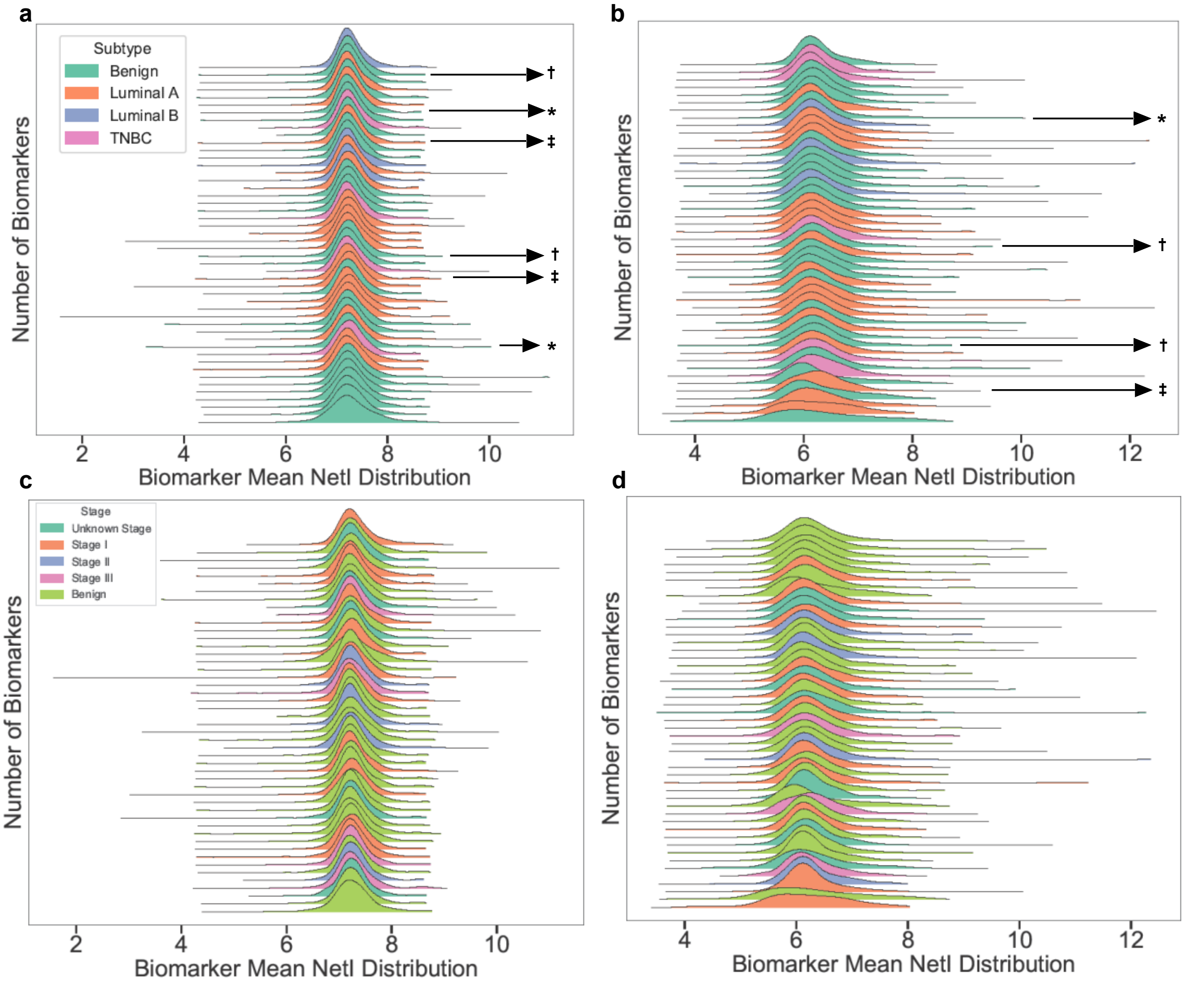

**SI Figure 5: Joy plots annotated for donor characteristics demonstrate no trends in total antibody distribution related to conversion to cancer status, prior cancer history, treatment status, or cancer stage.** NetI for both the **a,c)** IgM and **b,d)** IgG response to the 525 TAAs in the panel are plotted for each donor. The height of each curve represents the relative number of biomarkers at a given NetI value, and the width of each distribution represents the total range of each donor's antibody distribution. **(a, b)** Distributions are sorted by peak height, and color designates the diagnostic class and subtype of each donor as indicated in the legend in the top left panel **a**. An asterisk (*) identifies benign donors that developed breast cancer within two years following the time of sample collection, a dagger (†) identifies benign patients with a history of cancer, and a double dagger (‡) identifies donors receiving chemotherapy treatments at the time of sample collection. **(c, d)** Distributions are sorted in ascending order by distribution mean, and color designates the disease stage of each donor as indicated in the legend in the top left panel **c**.

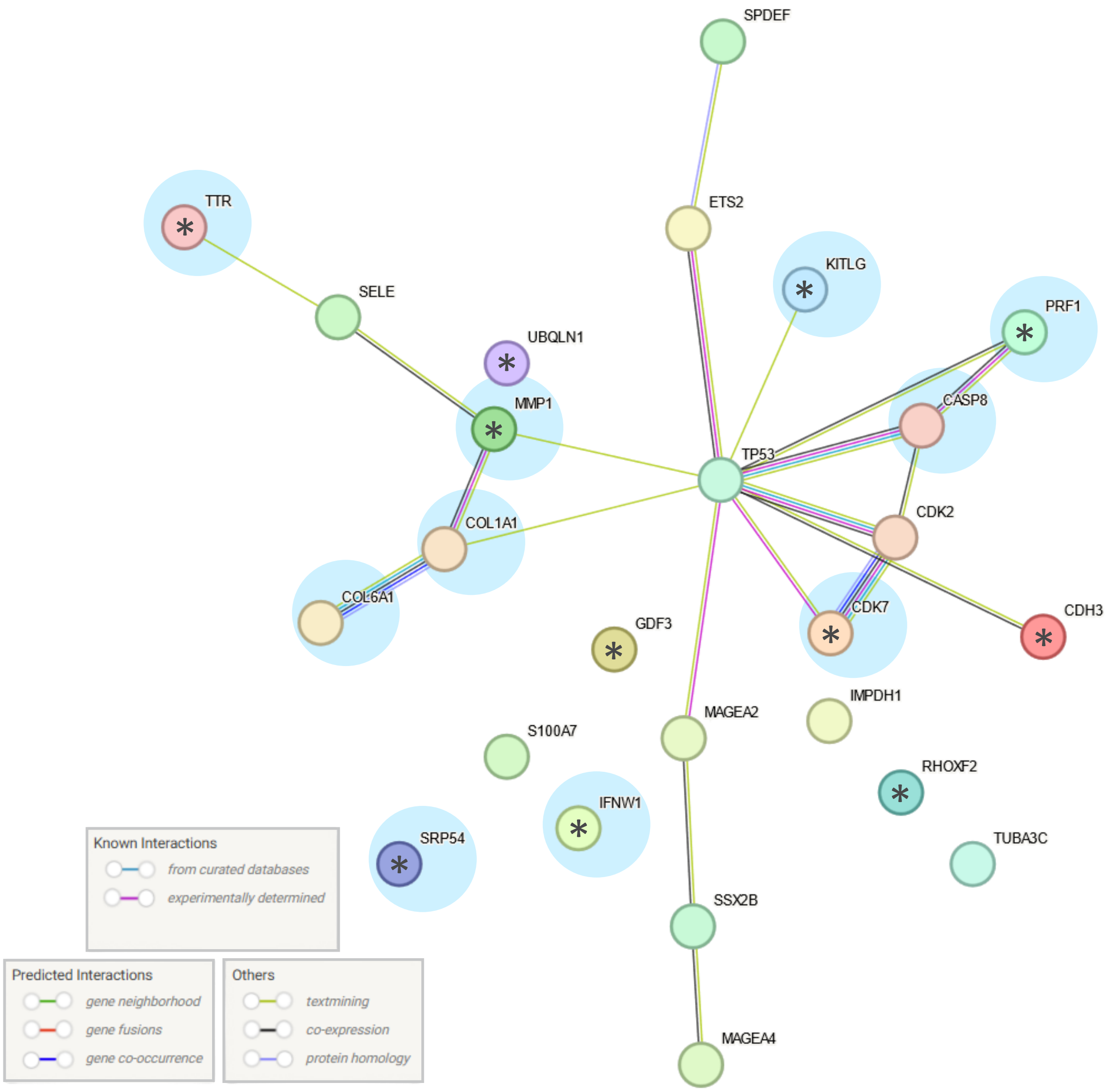

**SI Figure 6: Pathway analysis performed on antigens recognized by differentially expressed TAAbs identifies functional ties to TP53.** Each antigen is represented by a colored circle and labeled. A blue circle overlayed on top of an antigen indicates that the respective TAAb expression is elevated in the benign group rather than the cancer group. An asterisk (*) indicates that the antigen indicated has elevated TAAb expression in the IgM group rather than the IgG group. Different colored lines indicate different links between antigens as indicated by the legend at the bottom left.

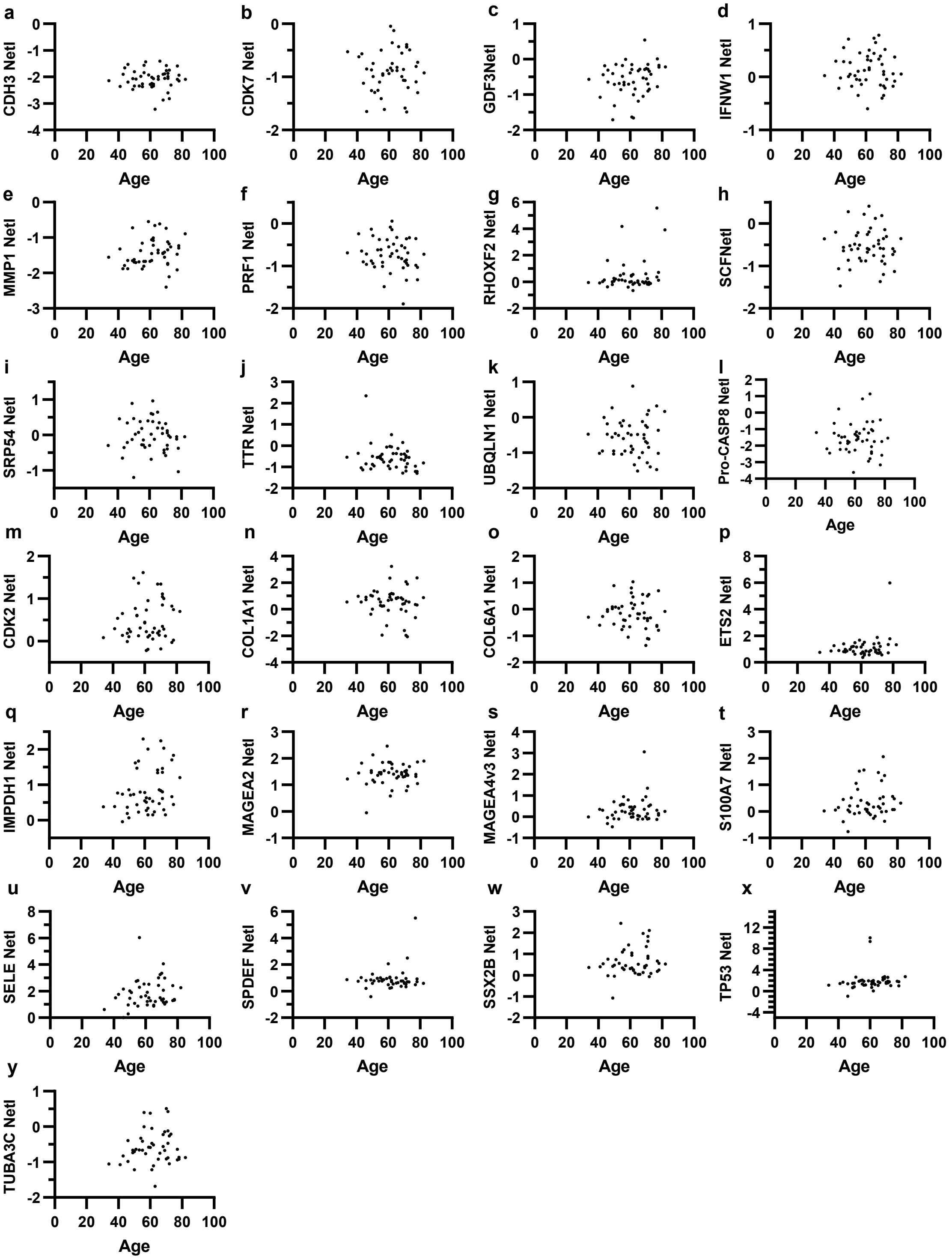

**SI Figure 7: No correlation is observed between age and antibody expression levels in the differentially expressed antibodies identified from the Normalized NetI data.** Each point represents an individual donor, with Normalized NetI values for each differentially expressed IgM (**a-k**) or IgG (**l-y**) antibody plotted against donor age. Normalized NetI is the average of four technical replicates (or two to three if any replicates were removed for a high CV > 20%).

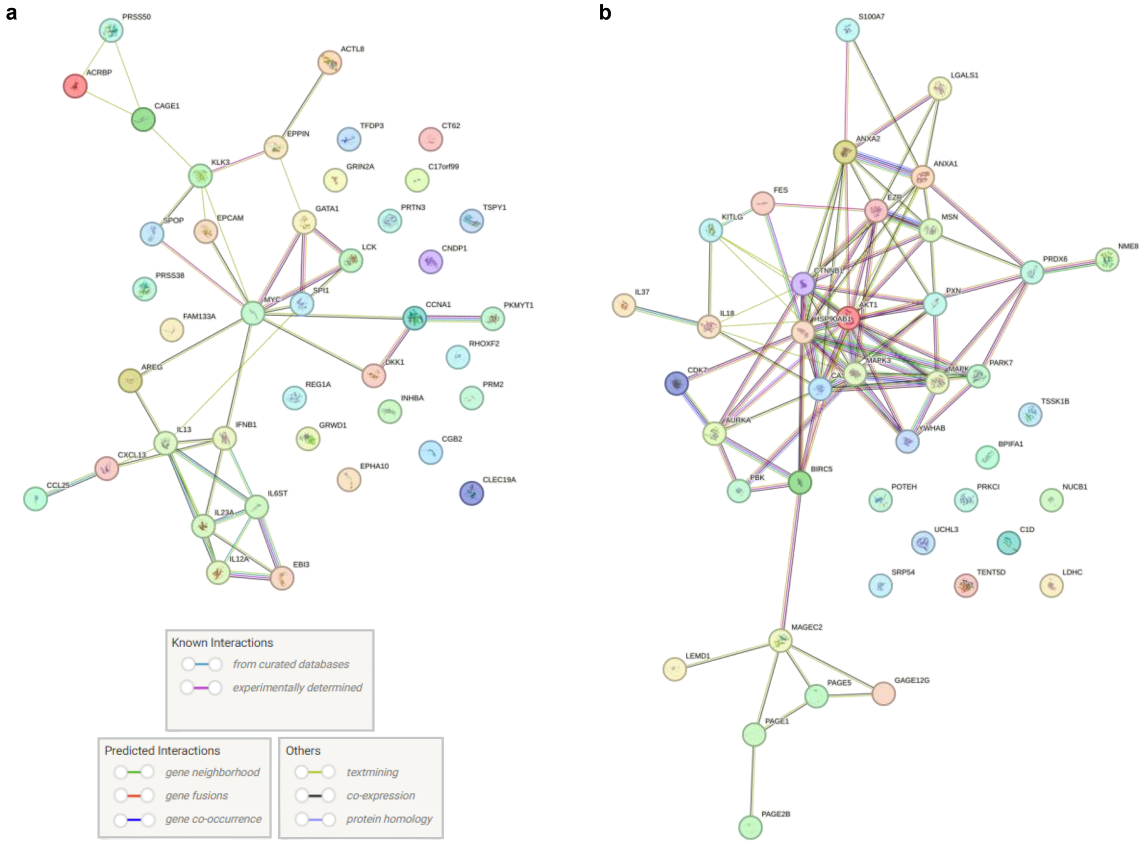

**SI Figure 8: Patient-level pathway analysis identifies unique personal immune profiles.** Pathway analysis for individual donors (**a**, **b**), with each antigen targeted by a high-titer IgG TAAb represented by a colored circle and labeled. Colored lines indicate different links between antigens as indicated by the legend at the bottom left.

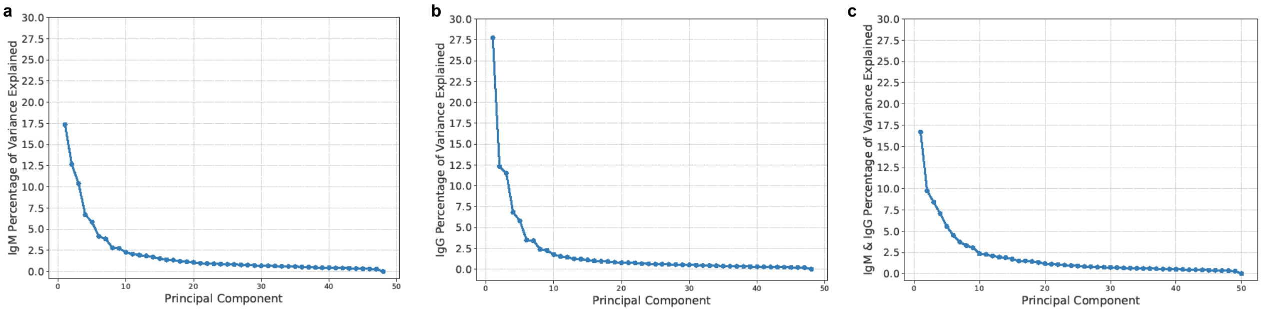

**SI Figure 9: Scree plots show limited variance captured by the first principal components in donors' antibody distributions.** The percentage of variance explained by each principal component is plotted in descending order across the x-axis for principal component analyses of the total **a)** IgM, **b)** IgG, and **c)** combined IgM and IgG Normalized NetI datasets.

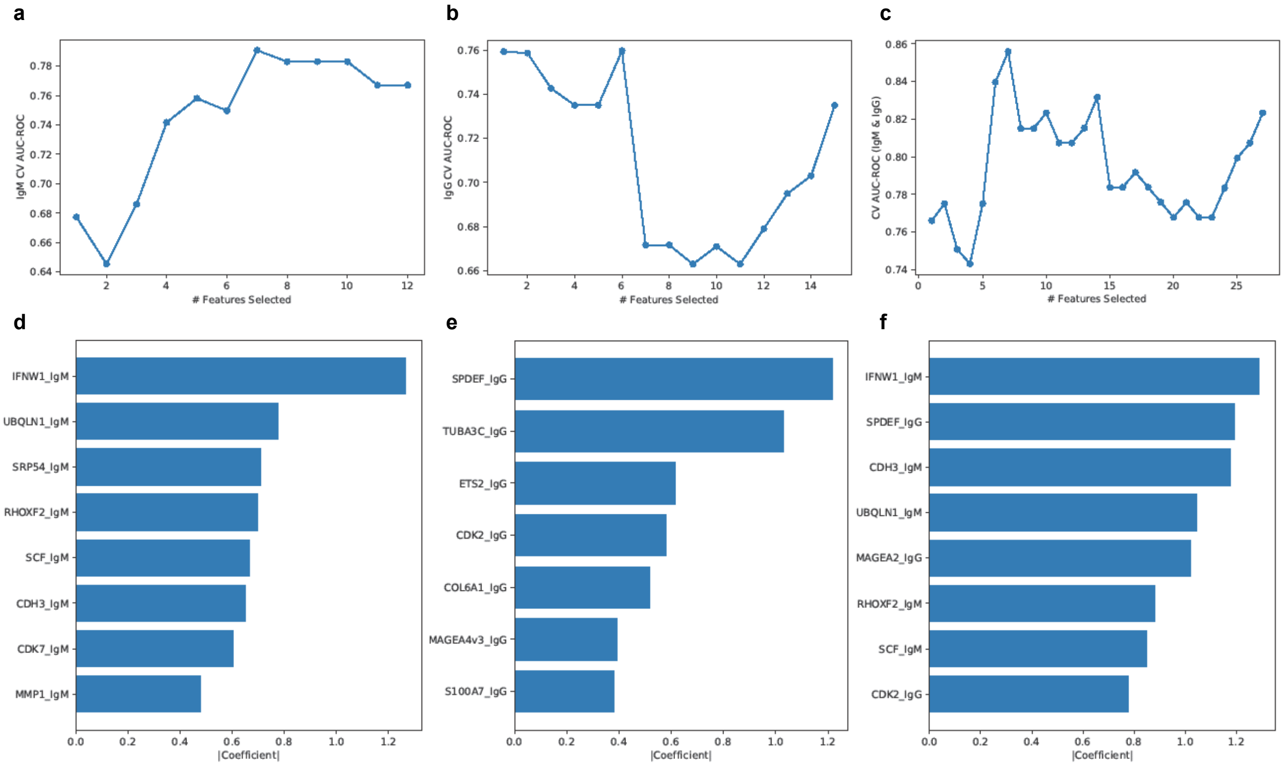

**SI Figure 10: Feature selection and feature importance plots inform the optimal number of features selected for classifier performance.** Feature selection plots show classifier AUC-ROC performance plotted against the number of features included in the **a)** IgM, **b)** IgG, and **c)** combined IgM and IgG classifiers. The features selected for each classifier are ranked by importance in their respective plots (**d-f**), with relative importance signified by the magnitude of the coefficient.
